# Relationship between alcohol consumption and dementia with Mendelian randomization approaches among older adults in the United States

**DOI:** 10.1101/2023.12.22.23300298

**Authors:** Kyle A. Campbell, Mingzhou Fu, Elizabeth MacDonald, Matthew Zawistowski, Kelly M. Bakulski, Erin B. Ware

**Affiliations:** Department of Epidemiology, University of Michigan School of Public Health, Ann Arbor, Michigan, USA; Department of Biostatistics, University of Michigan School of Public Health, Ann Arbor, Michigan, USA; Institute for Social Research, University of Michigan, Ann Arbor, Michigan, USA

**Author notes:** These authors contributed equally to this manuscript (Kelly M. Bakulski, Erin B. Ware). Corresponding author: Erin B. Ware, Population Neurodevelopment and Genetics (PNG), Institute for Social Research, Survey Research Center, 426 Thompson St. #3320, Ann Arbor, MI 48104.

**Keywords:** Dementia, cognitive impairment, alcohol, epidemiology, older adults, Mendelian randomization

## Abstract

**Background:** In observational studies, the association between alcohol consumption and dementia is mixed.

**Methods:** We performed two-sample Mendelian randomization (MR) using summary statistics from genome-wide association studies of weekly alcohol consumption and late-onset Alzheimer’s disease and one-sample MR in the Health and Retirement Study (HRS), wave 2012. Inverse variance weighted two-stage regression provided odds ratios of association between alcohol exposure and dementia or cognitively impaired, non-dementia relative to cognitively normal.

**Results:** Alcohol consumption was not associated with late-onset Alzheimer’s disease using two-sample MR (OR=1.15, 95% confidence interval (CI):[0.78, 1.72]). In HRS, doubling weekly alcohol consumption was not associated with dementia (African ancestries, n=1,322, OR=1.00, 95% CI [0.45, 2.25]; European ancestries, n=7,160, OR=1.37, 95% CI [0.53, 3.51]) or cognitively impaired, non-dementia (African ancestries, n=1,322, OR=1.17, 95% CI [0.69, 1.98]; European ancestries, n=7,160, OR=0.75, 95% CI [0.47, 1.22]).

**Conclusion:** Alcohol consumption was not associated with cognitively impaired, non-dementia or dementia status.

## 1. Introduction

Alzheimer’s disease and related dementias pose an increasing public health burden. The global prevalence of dementia in 2019 was 57.4 million and is projected to increase to 152.8 million by 2050 [1]. Identifying modifiable risk factors is critical. Alcohol consumption is a common exposure in the United States [2], and prior evidence is mixed regarding alcohol consumption as a risk factor for dementia. The recent Lancet Commission review on dementia prevention, intervention, and care concluded that evaluating previously published evidence examining alcohol consumption and dementia was “particularly challenging,” given alcohol’s complex relationship with other cultural and health factors [3]. Further research is needed to clarify the relationship between alcohol consumption and Alzheimer’s disease and related dementias to inform public health practices and reduce their incidence.

Low to moderate levels of alcohol consumption have been inconsistently associated with a reduced risk of dementias in systematic reviews and meta-analyses [4–7]. Several noted that publication bias of positive results should be considered [5, 7]. Operational definitions of alcohol consumption have varied [5, 7, 8]. A large dose-dependent meta-analysis found low alcohol consumption (≤12.5g/day) was protective against dementia risk [4]. Conversely, another dose-response meta-analysis of six prospective cohorts identified a linear association between alcohol consumption and risk of cognitive impairment, with an increased risk of progression to dementia with alcohol intake greater than 16 drinks per week (27.5g/day) [9]. The Lancet Commission review found in an updated meta-analysis of three studies that excessive alcohol consumption (>168 grams per week) in midlife was associated an 18% increase in dementia risk compared to lighter consumption [3]. A 2019 systematic scoping review on alcohol use and dementia concluded that a lack of control for confounding remains a challenge in the field [8]. Additional robust investigations are needed to better understand the relationship between alcohol consumption and dementia risk.

Genetic causal inference techniques such as Mendelian randomization may provide clearer evidence of a potential causal relationship between alcohol consumption and dementia. Mendelian randomization is a statistical method used to infer causality between a risk factor and outcome; it uses measured variation in genes as an instrumental variable for the risk factor of interest [10]. Mendelian randomization can overcome residual confounding and reverse causation typically associated with observational studies [10]. Prior Mendelian randomization investigations have largely found null associations between alcohol use and cognitive impairment or dementia [11–15]. Several of those studies only assessed cognitive impairment and relied on individual single nucleotide polymorphisms in the alcohol metabolism genes *ALDH2* and *ADH1B* as genetic instruments [11–13]. Only one such study of 235 rural Chinese individuals found an increased risk of cognitive impairment with increased alcohol consumption [12]. All studies were conducted in predominately Asian or European ancestries populations [11–15]. Further research in additional populations and genetic loci leveraging Mendelian randomization approaches may clarify the relationship between alcohol consumption and dementia risk.

In this study, we test the hypothesis that alcohol consumption is a risk factor for cognitively impaired non-dementia or a risk factor for dementia using three approaches. First, with two-sample Mendelian randomization, we use summary statistics from large genome-wide association studies to test the relationship between weekly alcohol consumption and late onset Alzheimer’s disease. Second, we perform a conventional cross-sectional analysis in the Health and Retirement Study (HRS), a large, multi-ethnic US cohort, to test the association between weekly alcohol consumption and dementia or cognitively impaired non-dementia. Third, we apply one-sample Mendelian randomization in the HRS using the same summary statistics to create genome-wide or instrument-based polygenic risk scores to test the association between weekly alcohol consumption and dementia or cognitively impaired non-dementia. We also perform robust single nucleotide polymorphisms (SNPs)-wise Mendelian randomization analyses. We therefore address critical limitations of previous observational and Mendelian randomization investigations by addressing the potential for residual confounding, reverse causation, and exposure assessment with Mendelian randomization methods alongside strong outcome assessment and sample diversity.

## 2. Methods

### 2.1 Summary statistics for alcohol consumption and late onset Alzheimer’s disease

To create a genetic instrument for alcohol consumption exposure, we use summary statistics from a large (n=941,280) publicly available genome-wide association meta-analysis of weekly alcohol consumption [16]. Briefly, effect sizes between individual single nucleotide polymorphisms (SNPs) and alcohol consumption as well as corresponding standard errors were generated within 34 individual cohorts with a standardized analysis plan, adjusted for age, age-squared, sex, and genetic ancestry principal components. Cohort characteristics were similar and of predominately European ancestries; non-European ancestry individuals were excluded from genetic analyses. The alcohol consumption phenotype was defined as log-transformed drinks per week among current drinkers as a continuous variable. If a study recorded binned response ranges, the midpoint of that range was used. SNPs present in only two or fewer studies or variants with minor allele frequency less than 0.001 were excluded. Association summary statistics were meta-analyzed with genomic controls using a novel fixed effects approach to account for study heterogeneity.

To create a genetic instrument for late onset Alzheimer’s disease, we use summary statistics from a large publicly available meta-analysis of late onset Alzheimer’s disease (n=21,982 cases, 41,944 controls) [17]. Briefly, the International Genomics of Alzheimer’s Project consortium meta-analyzed summary statistics from 46 case-controls studies from the following consortia: Alzheimer’s Disease Genetics Consortium, Cohorts for Heart and Aging Research in Genomic Epidemiology, European Alzheimer’s Disease Initiative, and Genetic and Environmental Risk in Alzheimer’s Disease/Defining Genetic, Polygenic and Environmental Risk for Alzheimer’s Disease Consortium. The outcome of interest was clinically diagnosed Alzheimer’s disease. Non-European ancestry individuals were excluded from analysis. Single variant additive genotype models adjusting for age, sex, and genetic ancestry principal components were used to identify SNPs associated with Alzheimer’s disease. Association summary statistics were meta-analyzed with inverse variance weighting and genomic control. Data for the outcome genome-wide association study was accessed through the MRC Integrative Epidemiology Unit Open GWAS Project database (accession number ieu-b-2) [18].

### 2.2 Health and Retirement Study sample

The Health and Retirement Study (HRS) is an ongoing longitudinal, nationally representative panel cohort of aging Americans over 50 years old and their partners that aims to capture health and demographic data surrounding retirement [19]. Since 1992, HRS has biennially collected data related to aging, including health and economic metrics at both the household and individual level from over 43,000 participants through mail-in surveys, telephone calls, leave-behind questionnaires, and face-to-face interviews. New participants are recruited every six years. Specifically, for each wave half of the participants are randomly assigned to the face-to-face method of data collection, which also includes collection of biomarkers, while the other half completes the core interview only through telephone interviews. For the subsequent wave collection two years later, the groups alternate between face-to-face and telephone interviews, ensuring that biomarker data is available at the individual level for all participants every four years. This study utilizes the 2012 cross-sectional wave of the HRS. Compared to other waves (with available genetic data for participants), the 2012 wave had the smallest number of missing values for cognitive status. HRS is administered by the Institute for Social Research at the University of Michigan and is funded by the National Institute on Aging (U01AG009740) and Social Security Administration. Informed consent was obtained from all participants. This secondary data analysis was approved by the University of Michigan Institutional Review Board (HUM00128220).

### 2.3 Genetic data

HRS face-to-face interviews between 2006 and 2012 included saliva collection for genotyping on the Illumina HumanOmni2.5 microarray platform by the Center for Inherited Disease Research. Genotype data was then analyzed by the Quality Assurance and Quality Control analysis team at the University of Washington and imputed to the 1000 Genomes Project reference panel [20]. The HRS uses principal component analysis on genome-wide SNPs to determine genetic ancestry [21]. The HRS released genetic data only on participants assigned European genetic ancestry and self-identified race/ethnicity as non-Hispanic White as well as assigned African genetic ancestry and self-identified race/ethnicity as non-Hispanic Black/African American [21]. To control for confounding due to population stratification, all analyses were conducted separately by ancestries and adjusted for five ancestry-specific principal components [21].

Polygenic risk scores are cumulative measurements of genome-wide genetic risk for a particular phenotype created by aggregating the effect of individual SNPs weighted according to effect size and allele dosage [22]. We used a publicly available polygenic risk score for weekly alcohol consumption computed in the HRS cohort [23]. The HRS polygenic risk score for an individual is calculated by summing the product of the weight (effect size from external meta-analysis [16]) and the allele dosage (0, 1, or 2) across all 1,399,824 SNPs overlapping the HRS and Liu et al., 2019 [16] weekly alcohol consumption GWAS meta-analysis (n=403,931). Due to data sharing limitations, the score was calculated without 23andMe data. We also created a polygenic risk score using the same approach among only SNPs that met instrument selection criteria (see **2.8 One-sample Mendelian randomization instrument selection and assessment** for further detail) using summary statistics from the exposure GWAS of weekly alcohol consumption (n=941,280) [16]. Polygenic risk scores were z-score standardized within ancestries for analysis.

### 2.4 Cognitive status outcome

Cognitive status of HRS participants was categorized as cognitively normal, cognitively impaired non-dementia, or dementia based on an additive 27-point score across multiple cognitive tests: an immediate and delayed 10-noun recall test, a backward count from 20 test, and a serial 7 subtraction test. Cut points corresponding to the three levels of cognitive status were based on the Langa-Weir classification, which has been clinically validated and has a sensitivity of 78% [24].

### 2.5 Alcohol consumption exposure

Alcohol consumption status was assessed through self-reported responses in the 2012 HRS wave. Participants were asked to provide the number of days per week that they consumed alcohol and the number of alcoholic drinks they consumed per day when they did drink alcohol. We calculated a continuous measure of weekly alcohol consumption by multiplying the number of days per week they reported drinking alcohol and the number of drinks consumed per day. We added one to this weekly alcohol consumption measure and applied a log2 transformation to reduce the effect of outliers and harmonize the exposure with the weekly alcohol consumption genome-wide association study [16]. When used as an exposure in a logistic regression model, the effect size for the resulting alcohol measure is interpreted as the odds ratio associated with a doubling in weekly alcohol consumption.

### 2.6 Covariates

We included potential confounders as covariates, including age (continuous: years), sex (categorical: male or female), educational attainment (categorical: no degree, high school diploma/GED/some college, or two year college degree or greater), ever smoking status (categorical: never smoking or ever smoking), current depressive symptoms (continuous: Center for Epidemiologic Studies Depression Scale), a health problems index corresponding to the number of comorbid conditions as communicated to the respondent by a doctor (continuous: 0-8 scale, 1 point each for any of: high blood pressure, diabetes, cancer, lung disease, heart disease, stroke, psychiatric problems, or stroke), marital status (categorical: married, separated/divorced, widowed, or never married), and retirement status (categorical: not retired, completely retired, partly retired, or irrelevant). Ever smoking status, depressive symptoms, the health index score, marital status, and retirement status variables were adapted from RAND HRS Longitudinal File 2020 (V1) produced by the RAND Center for the Study of Aging, with funding from the National Institute on Aging and the Social Security Administration. Santa Monica, CA (March 2023). The *APOE* gene is a risk factor for dementia [25]. There are three common *APOE* isoforms: ε2, ε3, and ε4 [26]. HRS participants were categorized by their *APOE* gene carrier status as ε2/ε2, ε2/ε3, ε2/ε4, ε3/ε3, ε3/ε4, and ε4/ε4 using genetic data imputed to the 1000 Genomes Project reference panel [20]. We therefore also included *APOE* ε4 allele status (categorical: 0, 1, or 2) as a precision variable.

### 2.7 Two-sample Mendelian randomization instrument selection and assessment

There was no sample overlap between the two meta-analyses from which we derived instrument-exposure and instrument-outcome summary statistics [16, 17]. Data preprocessing, harmonization, and analysis steps were performed using the *TwoSampleMR* R package (version 0.5.6) [27, 28]. Default settings were used unless otherwise noted. To ensure independence between SNPs, all potential index and secondary dependent SNPs (r^2^>0.001) were clumped at 10,000kb resolution using the European (EUR) super population reference panel from 1000Genomes [20]. We excluded SNPs that were not available in the reference EUR reference panel, which only contains biallelic SNPs with population minor allele frequency greater than 1% and excludes insertion-deletion mutations. If a particular exposure-linked SNP was not available in the outcome data, we substituted a secondary SNP in linkage disequilibrium (minimum r^2^>0.8) with the index SNP. Palindromic SNPs were inferred if minor allele frequency was less than 42%. We aligned alleles from the tagged SNP and inferred palindromic SNPs with minor allele frequency <30%. Finally, we harmonized exposure and outcome summary statistics according to the forward strand orientation; ambiguous strandedness or non-inferable palindromic SNPs were excluded. We assessed SNP instrument strength by its association with weekly alcohol consumption by calculating the proportion of variance explained and its corresponding F-statistic [29]. We assessed the potential for weak instrument bias by measuring SNP association strength using the proportion of variance explained [30] and the F-statistic [31].

### 2.8 One-sample Mendelian randomization instrument selection and assessment

One-sample Mendelian randomization two-stage least squares regression analyses in the HRS study sample were performed using the *MendelianRandomization* R package (version 0.5.6) [27, 28]. Polygenic risk score models were performed using the *ivtools* R package (version 2.3.0) [32]. Default settings were used unless otherwise noted. To ensure independence between SNPs, all potential index and secondary dependent SNPs (r^2^>0.001) were clumped at 10,000kb resolution using the EUR super population reference panel from 1000Genomes. We excluded SNPs that were not available in the reference EUR reference panel. If a particular exposure SNP was not available in the outcome data, we substituted a secondary SNP in linkage disequilibrium (minimum r^2^>0.8) with the index SNP. We included independent (r^2^<0.001) candidate exposure SNPs identified in the two-sample Mendelian randomization analysis that overlapped with the HRS study sample. We excluded SNPs with minor allele frequency less than 2%. We assessed exposure instrument strength using the F-statistic [31] derived from linear models. We also generated partial F-statistics in nested general linear models with or without the SNP present adjusted for principal components of genetic ancestry, additionally adjusted for demographic variables age, sex, and educational attainment, additionally adjusted for *APOE* ε4 allele status and potential confounders: ever smoking status, depressive symptoms, the comorbidities health index score, marital status, and retirement. As a sensitivity analysis, we separately assessed alcohol consumption polygenic risk scores as instrumental variables. To test the exclusion Mendelian randomization assumption, we regressed cognitive outcomes on each instrument, either individual SNPs or polygenic risk scores, using logistic models. Statistical significance indicating a potential violation of exclusion was assessed with 95% confidence intervals. To test the exogeneity assumption, we individually regressed each instrument, either SNPs or polygenic risk scores, on each potentially confounding covariate with general linear models and assessed the model F-statistic. Statistical significance indicating a potential violation of exogeneity was assessed at alpha=0.05.

### 2.9 Statistical analysis

We used a variety of two-sample Mendelian randomization estimators that are differentially robust to violations of Mendelian randomization pleiotropy assumptions, including inverse variance weighted, Egger regression [33], simple mode, and weighted mode Mendelian randomization estimates. To assess heterogeneity, we examined inverse variance weighted and Egger regression Q statistic estimates. To identify and assess outliers, we additionally performed pleiotropy residual sum and outlier Mendelian randomization (MR-PRESSO) and leave-one-out sensitivity analyses. To establish directionality between alcohol consumption and cognition outcomes, we used the Steiger test of directionality [27].

To assess the cross-sectional association between alcohol consumption and dementia or cognitively impaired non-dementia in the HRS sample, we performed conventional logistic regression in analyses using general linear models with a binomial link function in R (version 4.2.2). For the primary analysis, linear models were adjusted for all covariates as potential confounders or precision variables as described above. We also performed sensitivity analyses that tested unadjusted models, models adjusted for the demographic characteristics alone, and models adjusted for demographics and *APOE* ε4 allele status. All analyses were stratified by genetic ancestries.

We used a variety of one-sample Mendelian randomization estimators that are differentially robust to violations of Mendelian randomization pleiotropy assumptions, including inverse variance weighted, Egger regression [33], and weighted median Mendelian randomization estimates with two-stage least squares regression. For the primary analysis, as recommended in current best practices, Mendelian randomization models were adjusted for age, sex, and 5 principal components of genetic ancestry [34]. We also performed sensitivity analyses that tested unadjusted models, models adjusted for the demographic characteristics age, sex, and educational attainment alone, models adjusted for demographic characteristics and APOE ε4 allele status, and models adjusted for all potential precision variable and confounding variables as described above. All analyses were stratified by genetic ancestries and all models were minimally adjusted for 5 principal components of genetic ancestry. The alternatives to the inverse variance weighted approach are more robust to violations of the Mendelian randomization assumptions but come at the cost of statistical efficiency [35]. To assess heterogeneity, we examined inverse variance weighted and Egger regression Q statistic estimates. To identify and assess outliers, we additionally performed leave-one-out sensitivity analyses. We also tested the whole genome alcohol consumption polygenic risk score and the polygenic risk score constructed from only instrument-qualified SNPs as individual instruments in separate analyses using the two-stage least squares approach.

## 3. Results

### 3.1 Two-sample Mendelian randomization

The causal effect of alcohol consumption on cognition outcomes can be estimated using Mendelian Randomization if a genetic variant is associated with alcohol consumption (relevance), affects cognition outcomes only through its potential effect on alcohol consumption (exclusion), and is not confounded with cognition outcomes (exogeneity) (**Figure 1**). We first performed a two-sample Mendelian randomization analysis using summary statistics for 59 harmonized, genome-wide significant (p-value<1×10^−8^), and independent (r^2^<0.001) SNPs that met instrument selection criteria from large, publicly available meta-analyses of genome-wide association studies of weekly alcohol consumption (n=941,280) [16] and clinically diagnosed late onset Alzheimer’s disease (n=21,982 cases, 41,944 controls) [17] (**Supplementary Figure 1**).

**Figure 1.**
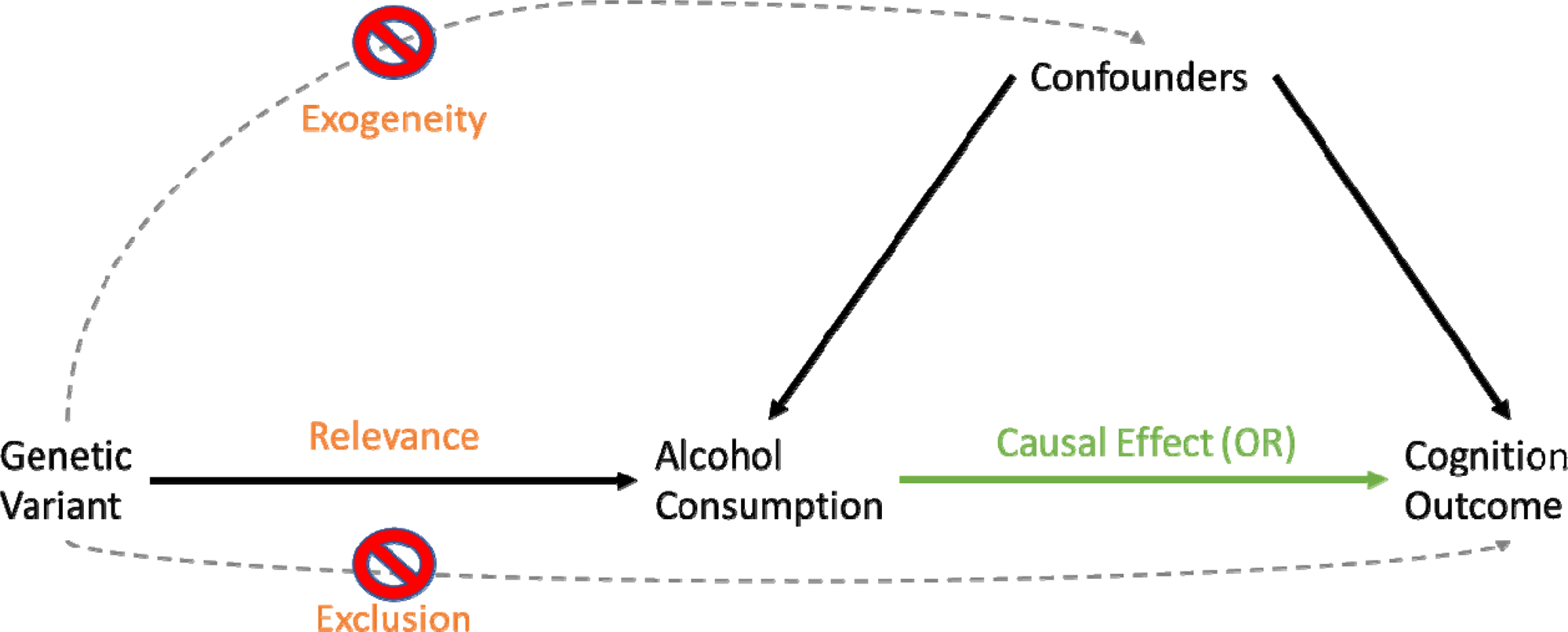
Mendelian randomization framework. Mendelian randomization framework presented as a directed acyclic graph. If a genetic variant is associated with alcohol consumption (Relevance), affects cognition outcomes only through its potential effect on alcohol consumption (Exclusion), and is not confounded with cognition outcomes (Exogeneity), we can estimate the causal effect of alcohol consumption on cognition outcomes.

The 59 SNPs that met instrument selection criteria had a median F-statistic of 37.80 (IQR=11.65) for the strength of the association between an individual genetic instrument and alcohol consumption. Of those, 7 SNPs were proxy SNPs from the Alzheimer’s disease GWAS in linkage disequilibrium (minimum r^2^>0.8) with SNP instruments from the alcohol consumption GWAS. rs1229984, a variant that encodes an *ADH1B* gene polymorphism that reduces liver alcohol clearance, had a particularly high F-statistic of 1,522.46. The Steiger test of causal directionality suggested that the hypothesis that alcohol consumption affects late onset Alzheimer’s disease for these SNPs was correct (p=1.64×10^−9^). The inverse variance weighted two-sample Mendelian randomization estimator revealed no evidence of a causal association between alcohol consumption and Alzheimer’s disease (OR=1.15, 95% CI [0.78, 1.72]) (**Table 1**). The rs1229984 SNP alone also revealed no evidence of an association (OR=0.90, 95% CI [0.57, 1.41]).

**Figure 2.**
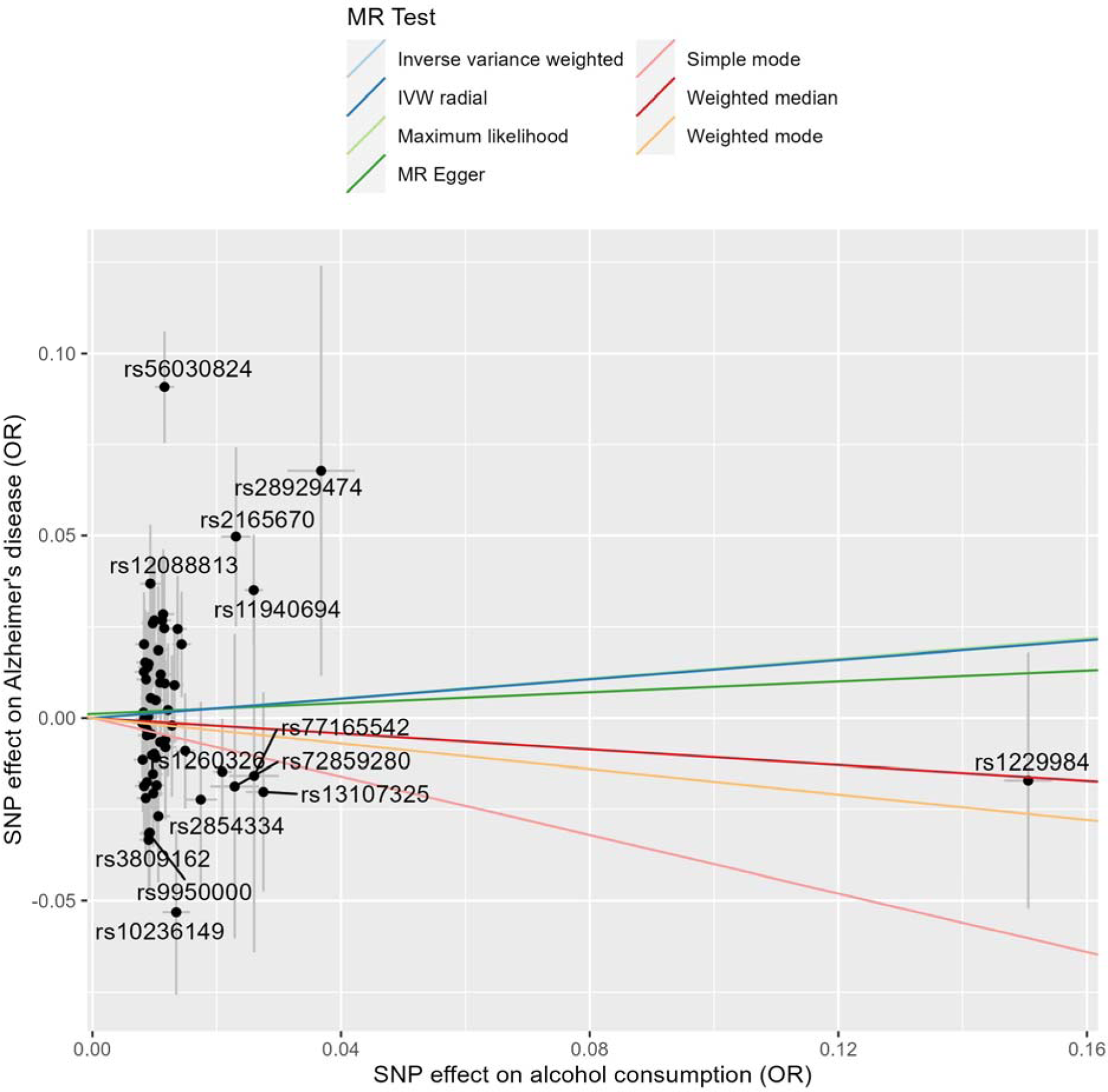
Primary two-sample Mendelian randomization test estimates for each individual single nucleotide polymorphism genetic instrument. The presented effect sizes are on the odds ratio (OR) scale. Grey lines represent the standard error for each single nucleotide polymorphism (SNP). Colored lines correspond to the relevant Mendelian randomization (MR) estimator.

**Table 1.** Two-sample Mendelian randomization estimates of previously published genome-wide association meta-analysis summary statistics of weekly alcohol consumption [16] and late onset Alzheimer’s disease [17] by test method.

| Estimator | Odds ratio | Lower 95% CI | Upper 95% CI | P-value |
| --- | --- | --- | --- | --- |
| Maximum likelihood | 1.15 | 0.86 | 1.53 | 0.35 |
| MR Egger | 1.08 | 0.57 | 2.03 | 0.82 |
| Weighted median | 0.90 | 0.59 | 1.38 | 0.62 |
| Inverse variance weighted | 1.14 | 0.77 | 1.70 | 0.51 |
| Simple mode | 0.67 | 0.18 | 2.52 | 0.56 |
| Weighted mode | 0.84 | 0.55 | 1.29 | 0.43 |

We next tested additional Mendelian randomization estimators that are more robust to horizontal pleiotropy, a violation of the exclusion assumption. Simple mode (OR=0.67, 95% CI [0.17, 2.61]), weighted mode (OR=0.84, 95% CI: [0.54, 1.32]), and weighted median (OR=0.90, 95% CI [0.59, 1.36]) estimators also indicated no evidence of an association. The Egger regression estimator similarly found no evidence of a causal association between alcohol consumption and Alzheimer’s disease (OR=1.08, 95% CI [0.57, 2.03]). The Egger regression intercept was not significantly different from 0, suggesting our results were not biased by directional horizontal pleiotropy (p=0.81).

Heterogeneity of effect estimates can indicate potential violations of Mendelian randomization or statistical model assumptions [36]. Cochrane’s Q statistic generated from the Egger regression (p=1.54×10^−5^) and inverse variance weighted (p=2.15×10^−5^) models suggest the presence of heterogeneity. However, in the presence of measurement error in the SNP-exposure association, the heterogeneity Q statistic is subject to type I error [36]. In such cases, exact modified weights should be used for global tests of heterogeneity [36]. Fixed effect (p=0.35) or random effects (p=0.57) exact weights indicated no evidence of heterogeneity.

To identify and assess outliers, we performed pleiotropy residual sum and (MR-PRESSO) Mendelian randomization and performed leave-one-out sensitivity analyses (**Supplementary Figure 2**). rs56030824 was consistently identified as an outlier across these approaches and was algorithmically determined to be an outlier in the MR-PRESSO model. The MR-PRESSO estimator before (p=0.51) or after (p=0.82) outlier removal provided no evidence of a casual association between alcohol consumption and Alzheimer’s disease. In summary, our two-sample Mendelian randomization analyses found no evidence of a potential causal association between alcohol consumption and Alzheimer’s disease.

### 3.2 Health and Retirement Study sample description

We included participants of African genetic ancestries (n=1,842) or European genetic ancestries (n=8,328) who were interviewed in the 2012 wave of the HRS. We excluded individuals who were missing genetic data, cognitive outcome, alcohol consumption, or covariate data (**Supplementary Figure 3**). All analyses were stratified by genetic ancestries. Cognitive status was associated with the exposure and confounders but not with genome-wide or instrument-based polygenic risk scores for alcohol consumption (**Table 2**).

**Table 2.**
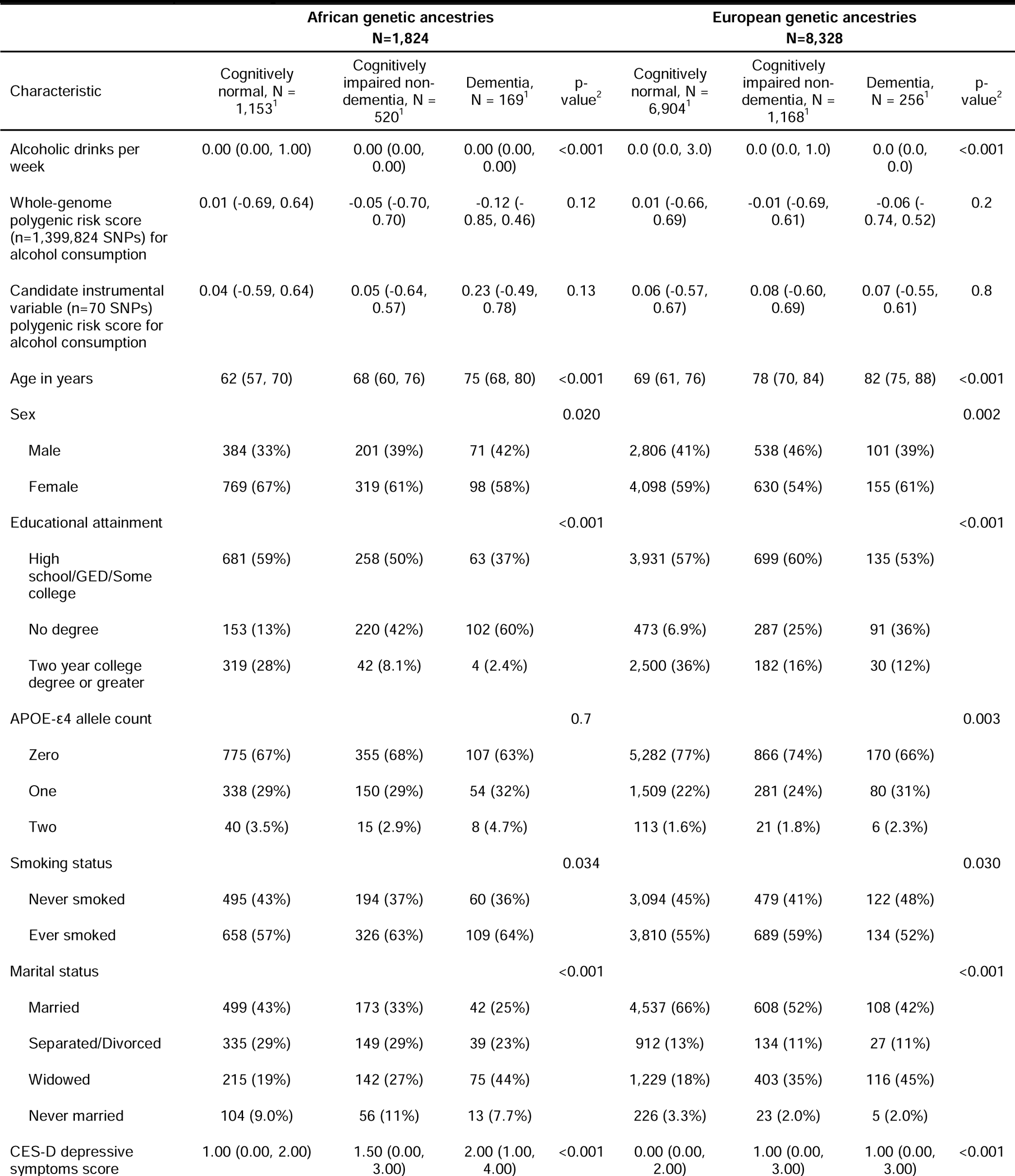
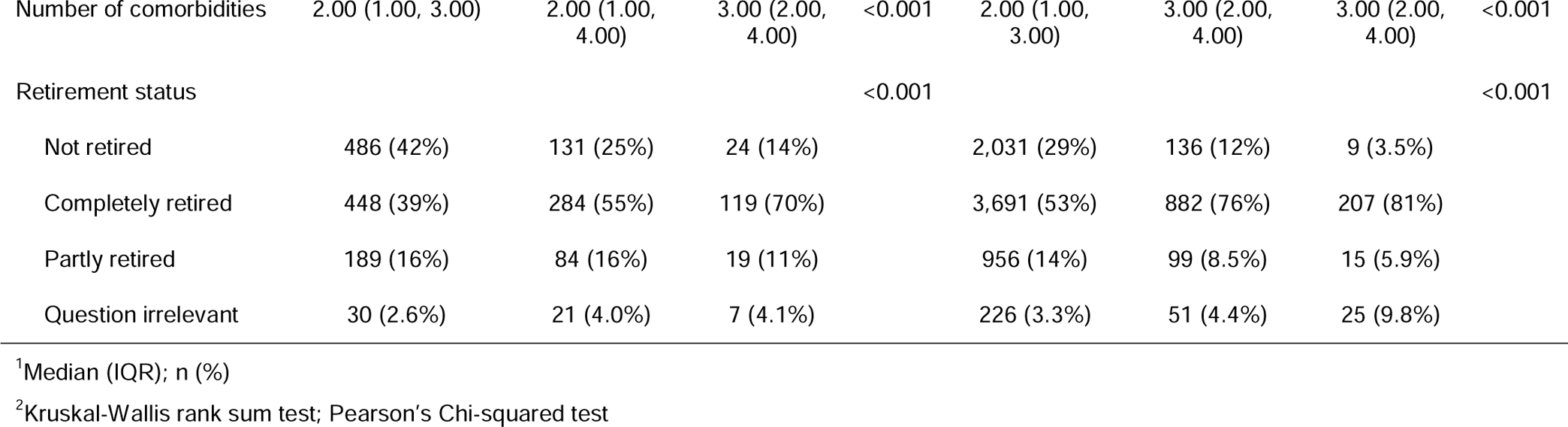
Bivariate table stratified by ancestries groups in the 2012 wave of the Health and Retirement study included in this empirical analysis.

### 3.3 Conventional logistic regression in the Health and Retirement Study

There was no evidence of an association between weekly alcohol consumption and dementia (OR=0.98, 95% CI [0.82, 1.17]) or cognitively impaired non-dementia (OR=0.96, 95% CI [0.82, 1.06]) in the African ancestries sample, in models adjusted for demographic variables age, sex, and educational attainment, *APOE* ε4 allele status, and potential confounders: ever smoking status, depressive symptoms, the comorbidities health index score, marital status, and retirement status (**Table 3**). In simpler models adjusted for only age, sex, educational attainment, and *APOE* ε4 allele status, we observed an apparent protective association between a doubling in alcoholic drink consumption and dementia (OR=0.60, 95% CI [0.50, 0.71]) as well as cognitively impaired non-dementia status (OR=0.87, 95% CI [0.83, 0.92]) (**Supplementary Table 1**); similar results were observed with even fewer covariates. A doubling in weekly alcohol consumption was inversely associated with dementia (OR=0.62, 95% CI [0.51, 0.73]) and cognitively impaired non-dementia (n=8,072, OR=0.89, 95% CI [0.84, 0.94]) in the European ancestries sample, in models adjusted for demographic variables age, sex, and educational attainment, *APOE* ε4 allele status, and additionally adjusted for potential confounders: ever smoking status, depressive symptoms, the comorbidities health index score, marital status, and retirement status (**Table 4**). This inverse association persisted in simpler models adjusted for fewer covariates (**Supplementary Table 2**).

**Table 3.**
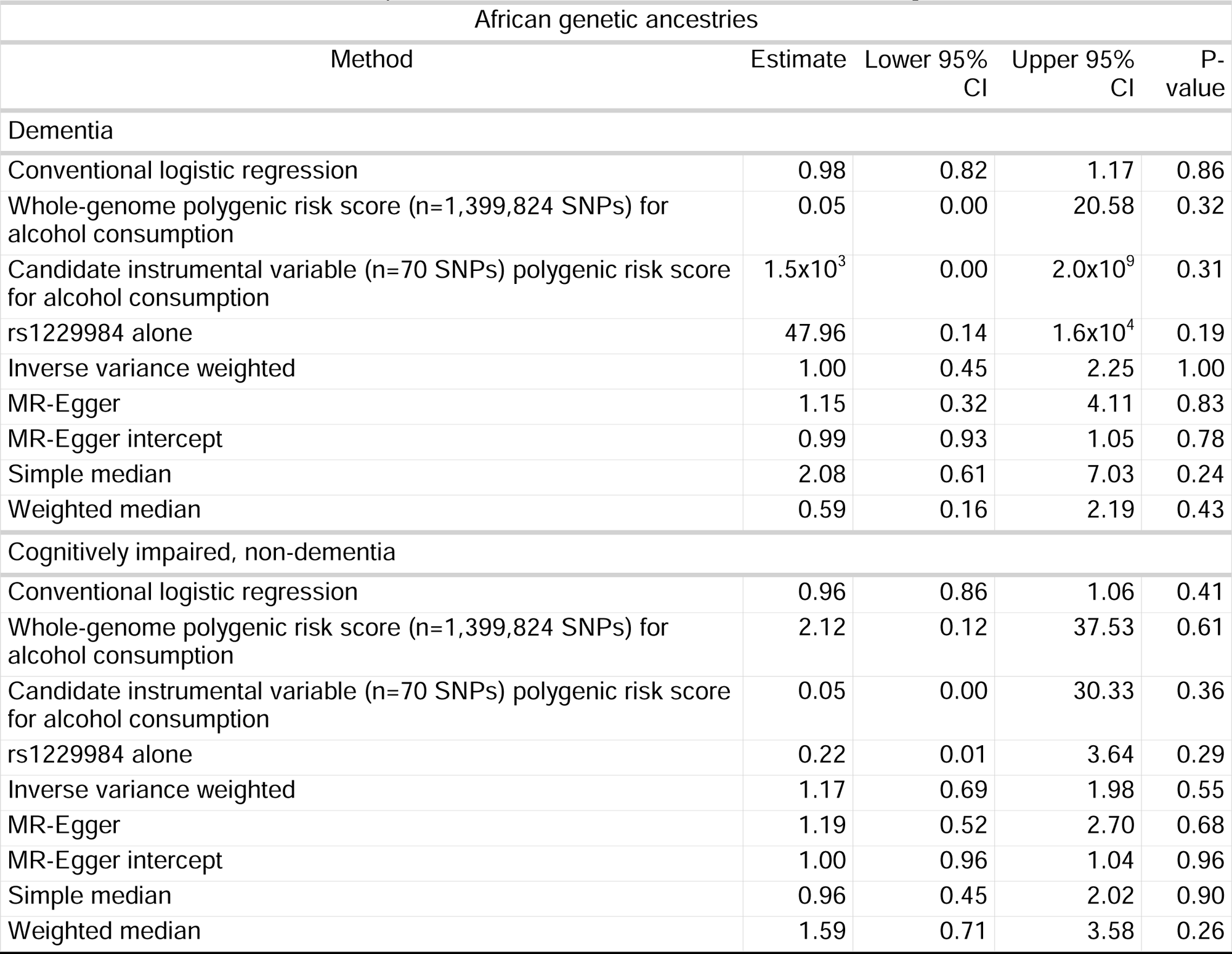
Table of effect estimates assessing the role of weekly alcoholic drinks consumption on cognitively impaired non-dementia or dementia status with conventional logistic regression or Mendelian randomization tests in the African ancestries sample (n=1,842) in the Health and Retirement Study.

| African genetic ancestries |  |  |  |  |
| --- | --- | --- | --- | --- |
| Method | Estimate | Lower 95% CI | Upper 95% CI | P-value |
| **Dementia** |  |  |  |  |
| Conventional logistic regression | 0.98 | 0.82 | 1.17 | 0.86 |
| Whole-genome polygenic risk score (n=1,399,824 SNPs) for alcohol consumption | 0.05 | 0.00 | 20.58 | 0.32 |
| Candidate instrumental variable (n=70 SNPs) polygenic risk score for alcohol consumption | $1.5 \times 10^3$ | 0.00 | $2.0 \times 10^9$ | 0.31 |
| rs1229984 alone | 47.96 | 0.14 | $1.6 \times 10^4$ | 0.19 |
| Inverse variance weighted | 1.00 | 0.45 | 2.25 | 1.00 |
| MR-Egger | 1.15 | 0.32 | 4.11 | 0.83 |
| MR-Egger intercept | 0.99 | 0.93 | 1.05 | 0.78 |
| Simple median | 2.08 | 0.61 | 7.03 | 0.24 |
| Weighted median | 0.59 | 0.16 | 2.19 | 0.43 |
| **Cognitively impaired, non-dementia** |  |  |  |  |
| Conventional logistic regression | 0.96 | 0.86 | 1.06 | 0.41 |
| Whole-genome polygenic risk score (n=1,399,824 SNPs) for alcohol consumption | 2.12 | 0.12 | 37.53 | 0.61 |
| Candidate instrumental variable (n=70 SNPs) polygenic risk score for alcohol consumption | 0.05 | 0.00 | 30.33 | 0.36 |
| rs1229984 alone | 0.22 | 0.01 | 3.64 | 0.29 |
| Inverse variance weighted | 1.17 | 0.69 | 1.98 | 0.55 |
| MR-Egger | 1.19 | 0.52 | 2.70 | 0.68 |
| MR-Egger intercept | 1.00 | 0.96 | 1.04 | 0.96 |
| Simple median | 0.96 | 0.45 | 2.02 | 0.90 |
| Weighted median | 1.59 | 0.71 | 3.58 | 0.26 |

**Table 4.** Table of effect estimates assessing the role of weekly alcoholic drinks consumption on cognitively impaired non-dementia or dementia status with conventional logistic regression or Mendelian randomization tests in the European ancestries sample (n=8,328) in the Health and Retirement Study.

| European genetic ancestries |  |  |  |  |
| --- | --- | --- | --- | --- |
| Method | Estimate | Lower 95% CI | Upper 95% CI | P-value |
| Dementia |  |  |  |  |
| Conventional logistic regression | 0.62 | 0.51 | 0.73 | 0.00 |
| Whole-genome polygenic risk score (n=1,399,824 SNPs) for alcohol consumption | 0.44 | 0.16 | 1.20 | 0.11 |
| Candidate instrumental variable (n=70 SNPs) polygenic risk score for alcohol consumption | 0.44 | 0.06 | 3.49 | 0.44 |
| rs1229984 alone | 2.50 | 0.14 | 45.59 | 0.54 |
| Inverse variance weighted | 1.37 | 0.53 | 3.51 | 0.52 |
| MR-Egger | 2.29 | 0.56 | 9.37 | 0.25 |
| MR-Egger intercept | 0.98 | 0.95 | 1.02 | 0.33 |
| Simple median | 0.45 | 0.11 | 1.88 | 0.27 |
| Weighted median | 1.89 | 0.44 | 8.20 | 0.40 |
| Cognitively impaired, non-dementia |  |  |  |  |
| Conventional logistic regression | 0.89 | 0.84 | 0.94 | 0.00 |
| Whole-genome polygenic risk score (n=1,399,824 SNPs) for alcohol consumption | 0.69 | 0.41 | 1.15 | 0.16 |
| Candidate instrumental variable (n=70 SNPs) polygenic risk score for alcohol consumption | 1.44 | 0.51 | 4.07 | 0.50 |
| rs1229984 alone | 3.31 | 0.87 | 12.54 | 0.08 |
| Inverse variance weighted | 0.75 | 0.47 | 1.22 | 0.25 |
| MR-Egger | 1.10 | 0.54 | 2.22 | 0.80 |
| MR-Egger intercept | 0.99 | 0.97 | 1.01 | 0.16 |
| Simple median | 0.58 | 0.28 | 1.20 | 0.14 |
| Weighted median | 0.90 | 0.44 | 1.84 | 0.77 |

### 3.4 One-sample Mendelian randomization in the Health and Retirement Study

Of 99 candidate drinks per week-associated SNPs, 70 harmonized, genome-wide significant, and independent SNPs with minor allele frequency greater than 2% were included for analysis (**Supplementary Figure 4**). Among African ancestries participants (n=1,842), the 70 SNPs that met instrument inclusion criteria had a mean partial F-statistic of 0.86 (SD: 1.07), indicating a potential violation of the relevance Mendelian randomization assumption, in models adjusted for age, sex, and genetic ancestry principal components. The whole genome (n=1,399,824 SNPs) and instrument-based (n=70 SNPs) polygenic risk scores had F-statistics of 2.26 and 1.12, respectively. Among European ancestries participants (n=8,328), the 70 SNPs that met instrument inclusion criteria had a mean partial F-statistic of 1.18 (SD:2.06) in age and sex-adjusted models (**Supplementary Figure 5**). The whole genome (n=1,399,824 SNPs) and instrument-based (n=70 SNPs) polygenic risk scores had F-statistics of 69.55 and 18.71, respectively. Results were generally robust to other model specifications. Exclusion assumption testing revealed that instruments were not associated with cognitive outcomes after false discovery adjustment. The whole genome alcohol consumption polygenic risk score was nominally associated with dementia in the African ancestries samples, whereas rs1229984 was nominally associated with cognitively impaired non-dementia within the European ancestries sample (**Supplementary Figure 6**). As expected, exogeneity assumption testing revealed consistent associations between principal components of genetic ancestry and genetic instruments. After false discovery p-value adjustment, we observed associations between 2 instruments (rs2854334, rs4548913) and educational attainment and rs10236149 and ever smoking status within the African ancestries sample (**Supplementary Figure 7**). We also observed associations between whole genome alcohol consumption polygenic risk score and ever smoking status, rs10506274 and marital status, rs1123285 and *APOE*-ε4 status, and 3 instruments (rs1229984, rs28680958, and our alcohol consumption PGS) and educational attainment in the European ancestries sample (**Supplementary Figure 8**).

The age and sex-adjusted inverse variance weighted one-sample Mendelian randomization estimator revealed no evidence of a causal association between alcohol consumption and dementia among the African ancestries (n=1,322, OR=1.00, 95% CI [0.45, 2.25]) (**Table 3**) or European ancestries (n=7,160, OR=1.37, 95% CI [0.53, 3.51]) samples (**Table 4**). The age and sex-adjusted inverse variance weighted one-sample Mendelian randomization estimator also revealed no evidence of a causal association between alcohol consumption and cognitively impaired non-dementia among African ancestries (n=1,673, OR=1.17, 95% CI [0.69, 1.98]) or European ancestries (n=8,072, OR=0.75, 95% CI [0.47, 1.22]) samples. Heterogeneity of effect estimates can indicate potential violations of Mendelian randomization or statistical model assumptions [36]. Egger regression intercept estimates revealed no evidence of directional horizontal pleiotropy. Cochrane’s Q statistic generated from the Egger regression and inverse variance weighted models revealed no evidence of heterogeneity. Sensitivity analysis estimators were similar to the inverse variance weighted analysis. Several model estimates exhibited excessively high standard errors should be interpreted with caution considering the wide confidence interval and a lack of replication among alternative estimators. Our results were also robust to different model covariate specifications in both African ancestries (**Supplementary Table 1**) and European ancestries samples (**Supplementary Table 2**). Examination of leave-one-out sensitivity analyses revealed rs1229984 as an outlier, though this did not change the result of the hypothesis test using the inverse variance weighted approach (**Supplementary Figure 9**, **Supplementary Figure 10**, **Supplementary Figure 11**, **Supplementary Figure 12**).

Because rs1229984 was an outlier in the SNP-level instrument analysis and has a known biological connection to alcohol consumption, we tested rs1229984 as an individual instrument alongside the polygenic risk scores. Neither the whole genome polygenic risk score, instrument-based polygenic risk score, nor rs1229984 revealed evidence of a causal association between alcohol consumption and dementia or cognitively impaired non-dementia in either ancestries sample (Error! Reference source not found., Error! Reference source not found.). Likely due to a rare minor allele frequency of 4.7%, standard error model estimates for rs1229984 were high. A poor instrument relevance criterion for the African ancestries sample led to excessively high model standard errors, likely due to poor correspondence with the genetic background of the discovery genome-wide association study. Consequently, these results should be interpreted with caution.

## 4. Discussion

In this study, we tested the hypothesis that alcohol consumption is associated with dementia or cognitively impaired non-dementia using three complementary approaches. We first conducted a two-sample summary data-based Mendelian randomization analysis of large meta-analyses of genome-wide association studies of alcohol consumption (n=941,280) and late onset Alzheimer’s disease (n=21,982 cases, 41,944 controls). Second, we performed an empirical genetic ancestries-stratified (African ancestries or European ancestries) one-sample summary data-based Mendelian randomization analysis in the HRS. Third, we completed a conventional cross-sectional analysis in the same HRS samples. In the cross-sectional European HRS sample, we observed a strong protective association between a doubling in weekly alcohol consumption and decreased odds of dementia (n=8,072, OR=0.62, 95% CI [0.51, 0.73]) and cognitively impaired non-dementia (n=7,160, OR=0.89, 95% CI [0.84, 0.94]) compared to cognitively normal individuals. In contrast, both Mendelian randomization analyses, which are more robust to residual confounding and reverse causation, almost uniformly revealed no evidence of a causal association between alcohol consumption and dementia or cognitively impaired non-dementia. The inverse variance weighted estimator in the two-sample Mendelian randomization analysis revealed no evidence of an association between alcohol consumption and late onset Alzheimer’s disease (OR=1.15, 95% CI [0.78, 1.72]). Similarly, the one-sample inverse variance weighted Mendelian randomization estimator revealed no evidence of a causal association between alcohol consumption and dementia or cognitively impaired non-dementia in either ancestries sample. Our results leverage robust causal inference-informed genetic techniques in diverse populations to add to the emerging evidence that observational protective associations between alcohol consumption and dementia or cognitively impaired non-dementia risk are likely biased by residual confounding and reverse causation.

Our cross-sectional European ancestries sample observational results were consistent with previous reports of a protective association between increased alcohol consumption and a decreased risk of cognitive impairment and dementia. A systematic review in 2008 of 23 studies found that low to moderate alcohol use was associated with a 32-38% reduced risk of dementia compared to non-drinkers [5]. We observed a similar result in the European ancestries sample (OR=0.62, 95% CI[0.51, 0.73]). Our study addresses several key limitations in investigating the relationship between alcohol consumption and cognitive impairment or dementia. First, our Mendelian randomization satisfied required assumptions and should therefore be less vulnerable to confounding and reverse causation [10]. Second, Mendelian randomization results might better capture lifetime alcohol consumption exposure than midlife observational cohorts typically used to study this research question, including the HRS data we analyzed [37]. Third, the well-characterized HRS contains validated cognitive outcomes [24]. Finally, we report stratified analyses in both African and European genetic ancestries samples. However, the external weekly alcohol consumption GWAS we used to identify candidate instruments and construct polygenic risk scores [16] was a predominately European genetic ancestries sample, potentially explaining the poor satisfaction of the Mendelian randomization relevance assumption observed in our African genetic ancestries sample analysis. The execution of a simultaneous conventional cross-sectional analysis and Mendelian randomization analysis in the same European and African ancestries samples highlights the potential limitations of observational studies of alcohol and cognitive impairment or dementia risk.

Contrary to prior observational research, Mendelian randomization studies have predominately found null and sometimes harmful associations between alcohol consumption and cognitive impairment or dementia. A large biobank-based Mendelian randomization analysis of *ALDH2* at the SNP rs671 conducted among southern Chinese men above the age of 50 found no association between alcohol consumption and cognition [11]. A recent small, cross-sectional Mendelian randomization analysis of *ALDH2* rs671 in rural China revealed an association between alcohol consumption and mild cognitive impairment [12]. A large cohort study Mendelian randomization analysis of *ADH1B* single nucleotide polymorphism rs1229984 conducted among community-dwelling Australian men above the age of 65 found no association between alcohol consumption and mild cognitive impairment [13]. A two-sample Mendelian randomization study found no evidence of a causal relationship between alcohol consumption and late-onset Alzheimer’s disease; they did, however, find evidence of an association between alcohol consumption and earlier Alzheimer’s age of onset [14]. A polygenic risk score based two-sample Mendelian randomization analysis of many putative modifiable Alzheimer’s disease risk factors and Alzheimer’s disease phenotypes found no association between alcohol consumption and Alzheimer’s disease [15]. Our Mendelian randomization analyses were also predominantly null. The simple median estimator identified an association between a doubling in weekly alcohol consumption and dementia in the African ancestries sample (OR=3.56, 95% CI [1.05, 12.04]) Though, these results should be interpreted with caution because of likely violation of the relevance assumption for the African ancestries sample. Taken together, Mendelian randomization analyses consistently identify alcohol consumption as a null or harmful risk factor in the incidence of cognitive impairment or dementia.

There were several strengths to our study. We leveraged large, publicly available meta-analyses of genome-wide association studies of alcohol consumption and Alzheimer’s disease to perform a two-sample Mendelian randomization study with a variety of robust sensitivity analyses. We similarly conducted a robust one-sample Mendelian randomization and simultaneous cross-sectional analysis in large, diverse samples in the HRS with validated outcome assessments and detailed covariate data. We also selected instruments in the one-sample Mendelian randomization from the large, independent genome-wide association study of alcohol consumption. The use of Mendelian randomization approaches specifically address several sources of potential bias outlined as key limitations of the evidence base for this research question, including residual confounding, exposure assessment, and reverse causation [8]. A Mendelian randomization analysis of alcohol consumption may better represent lifetime exposure compared to self-reported drinking [38, 39].

There were also several limitations to our study. The cross-sectional analysis in our empirical samples in the HRS is subject to several sources of potential bias, including self-reported exposure assessment measurement bias, reverse causation, and selection bias issues related to competing risks of death for alcohol and dementia-related mortality. Future investigations may prioritize longitudinal approaches alongside advanced methods to account for sources of selection bias. Several one-sample Mendelian randomization models in the African ancestries sample had excessively high standard errors in parameter estimates. These high standard errors were likely from low minor allele frequency in certain genetic variants and the lack of generalizability of alcohol-instrument associations generated in non-African ancestries samples resulting in low relevance. More diverse genetic association studies in individuals of African descent should be prioritized to increase the feasibility of Mendelian randomization analyses in African and other non-European ancestries populations.

In summary, we tested the potential causal relationship between alcohol consumption and cognitive impairment or dementia using a conventional cross-sectional analysis in the 2012 wave of the HRS, stratified by European and African ancestries. We conducted two types of Mendelian randomization analyses: one in the same HRS samples and another independent two-sample Mendelian randomization analysis in large, publicly available genome-wide association study meta-analyses of weekly alcohol consumption and late onset Alzheimer’s disease. The HRS cross-sectional analysis replicated previously observed protective, if paradoxical, associations between a doubling in weekly alcohol consumption in a retirement age United States population of European ancestries and a decreased odds of cognitively impaired non-dementia (n=8,072, OR=0.89, 95% CI [0.84, 0.94]) or dementia (OR=0.62, 95% CI [0.51, 0.73]). We did not observe an association between a doubling in weekly alcohol consumption in a retirement age United States population of African ancestries and cognitively impaired non-dementia (OR=0.96, 95% CI [0.82, 1.06]) or dementia (OR=0.98, 95% CI [0.82, 1.17]). A one-sample Mendelian randomization analysis in the HRS found no association between alcohol consumption and cognitive outcomes investigated. We did not observe evidence of a relationship between alcohol consumption and dementia using the inverse variance weighted two-sample Mendelian randomization estimator (OR=1.15, 95% confidence interval (CI): [0.78, 1.72]), nor with complementary sensitivity analysis estimators. Taken together, our results suggest observational studies investigating the relationship between alcohol consumption and cognitive impairment or dementia may be affected by sources of systematic bias that may be at least partially addressed by Mendelian randomization approaches, including issues of residual confounding, reverse causation, and exposure assessment. We conclude that alcohol consumption should not be considered a protective exposure against cognitive impairment or dementia.

## Data Availability

The Health and Retirement Study genotyping data that supports the findings of this study are available in The National Institute on Aging Genetics of Alzheimer’s Disease Data Storage Site (accession number NG00119). Other Health and Retirement Study data products, including the exposure, outcome, and covariate data used in this study can be found at https://hrs.isr.umich.edu/data-products.

## Code Availability

All scripts to perform preprocessing and analysis are available (https://github.com/bakulskilab).

## Supplementary Figures and Tables

**Supplementary Figure 1.**
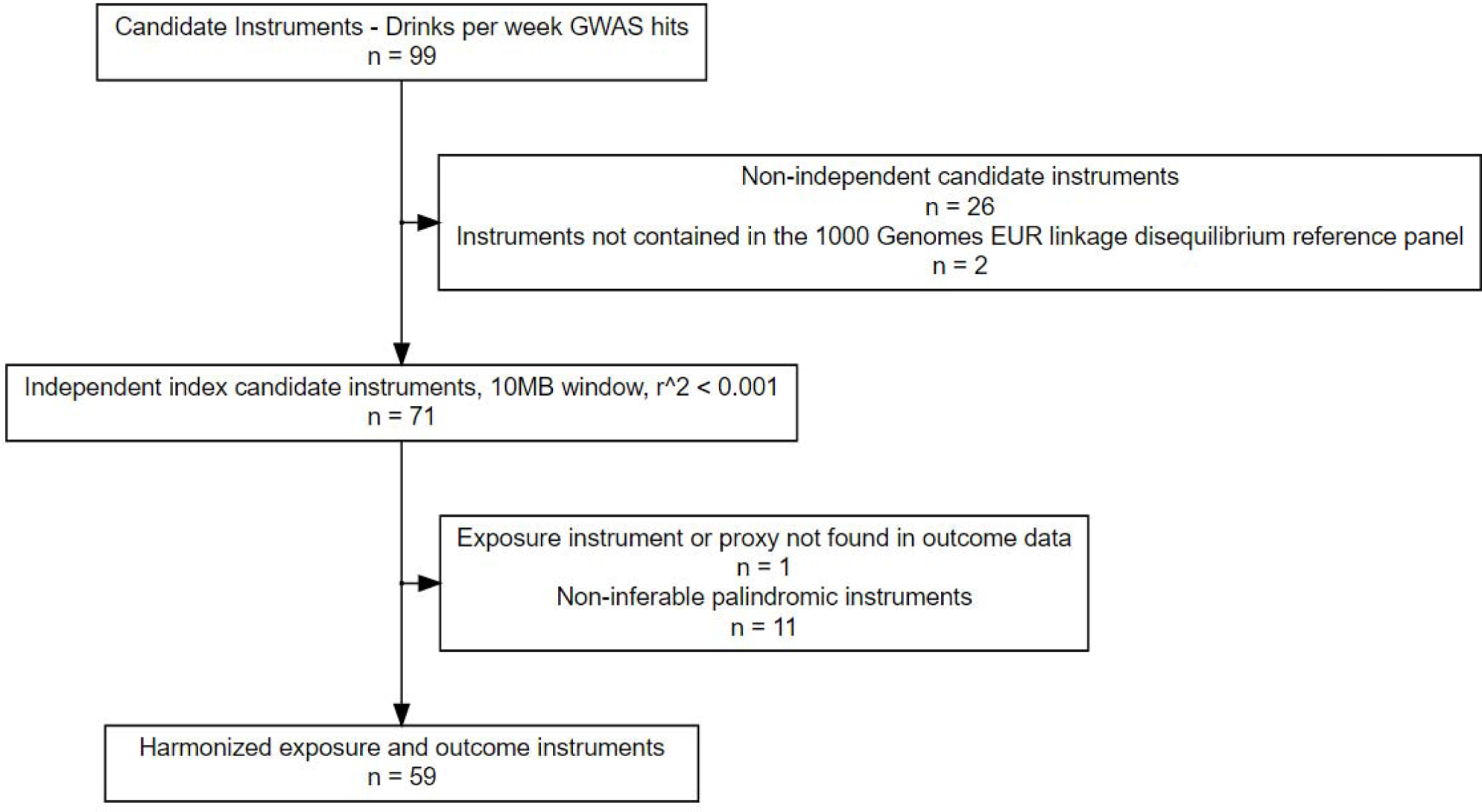
Inclusion-exclusion flowchart for two-sample Mendelian randomization single nucleotide polymorphism genetic instruments.

**Supplementary Figure 2.**
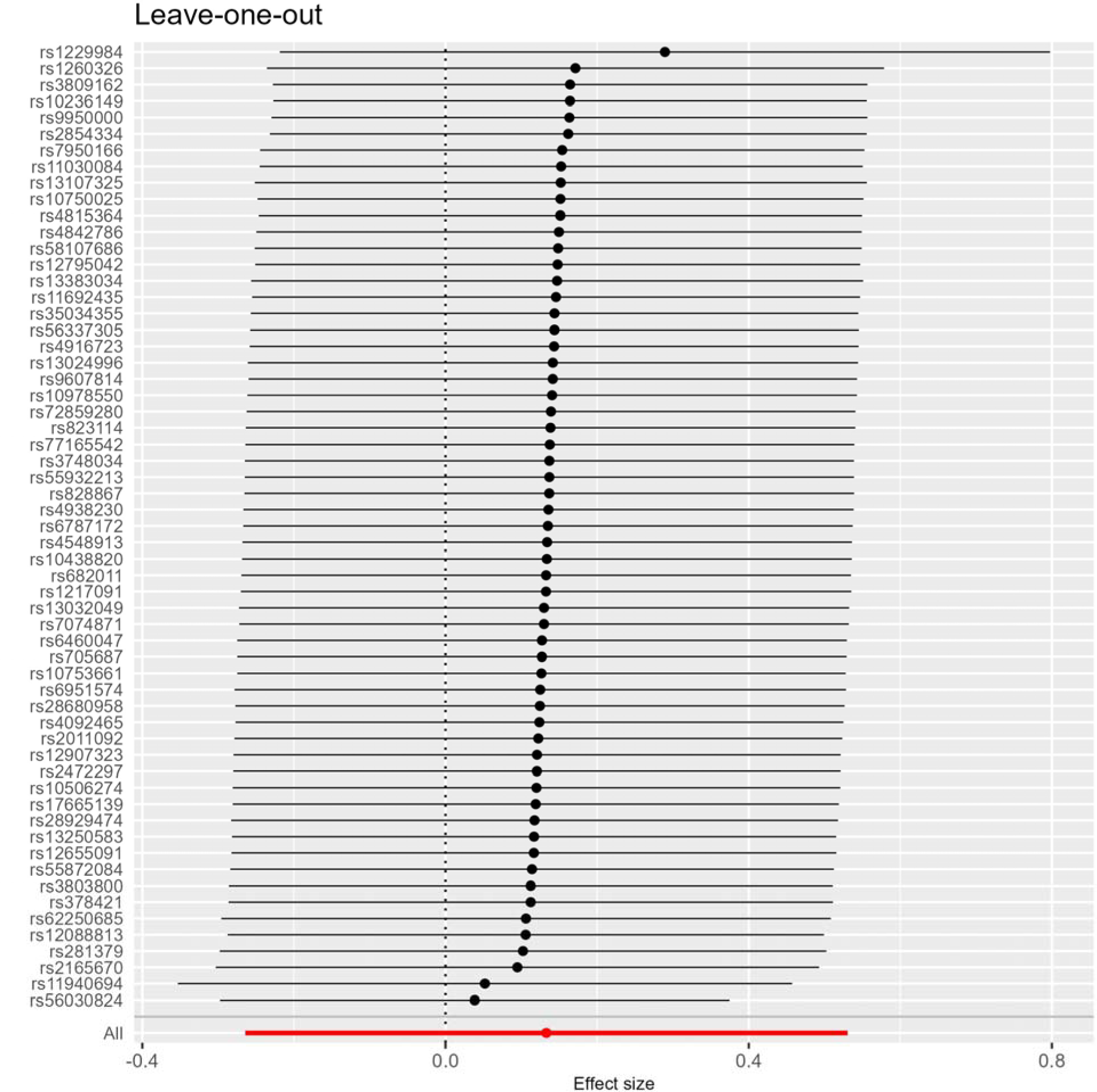
Leave-one-out sensitivity analysis for the inverse variance weighted two-sample Mendelian randomization estimator.

**Supplementary Figure 3.**
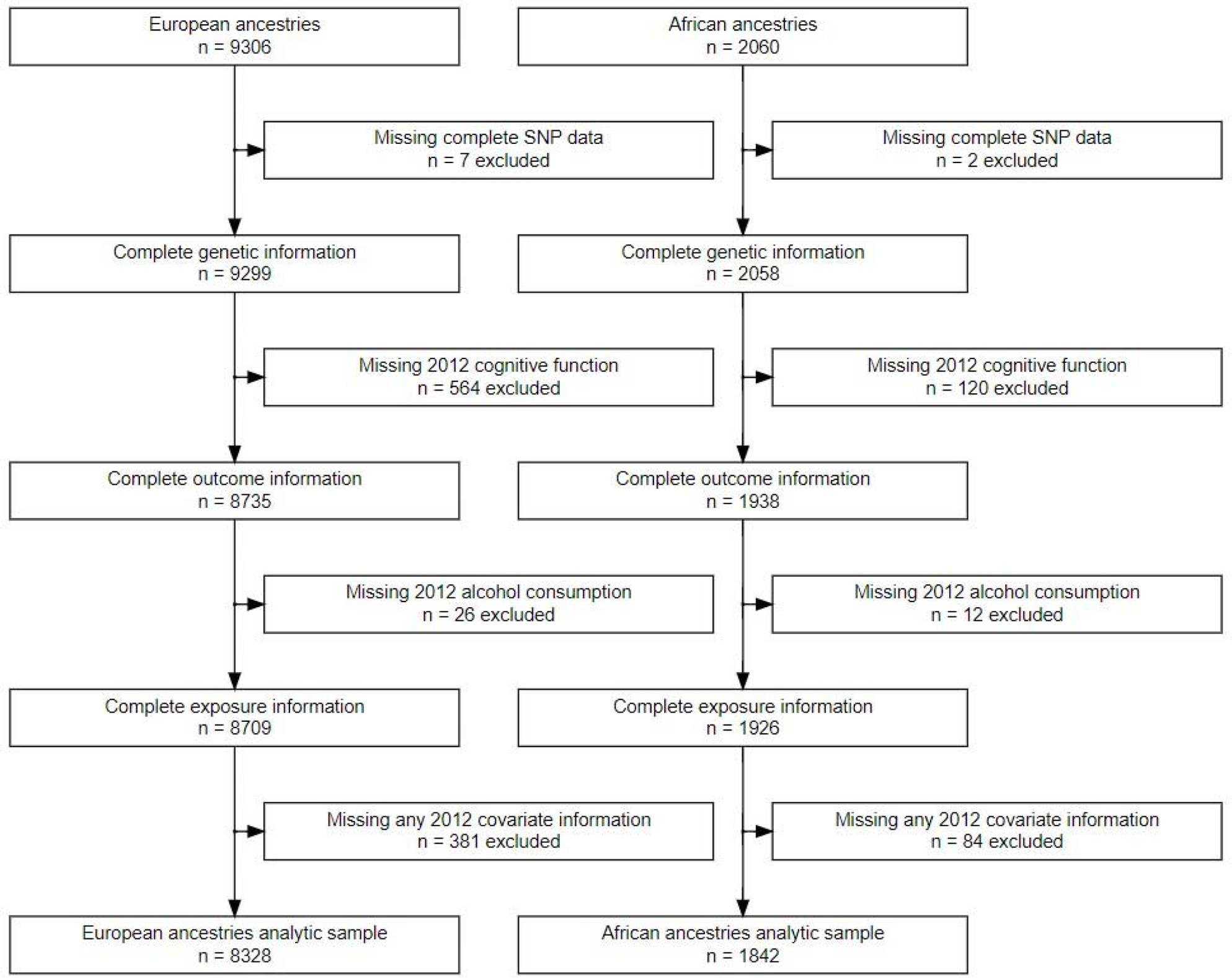
Inclusion-exclusion flowchart stratified by genetic ancestries for the empirical analysis in the 2012 wave of the Health and Retirement Study. The starting population included individuals with concordant broad-scale genetic ancestry and self-reported race and ethnicity.

**Supplementary Figure 4.**
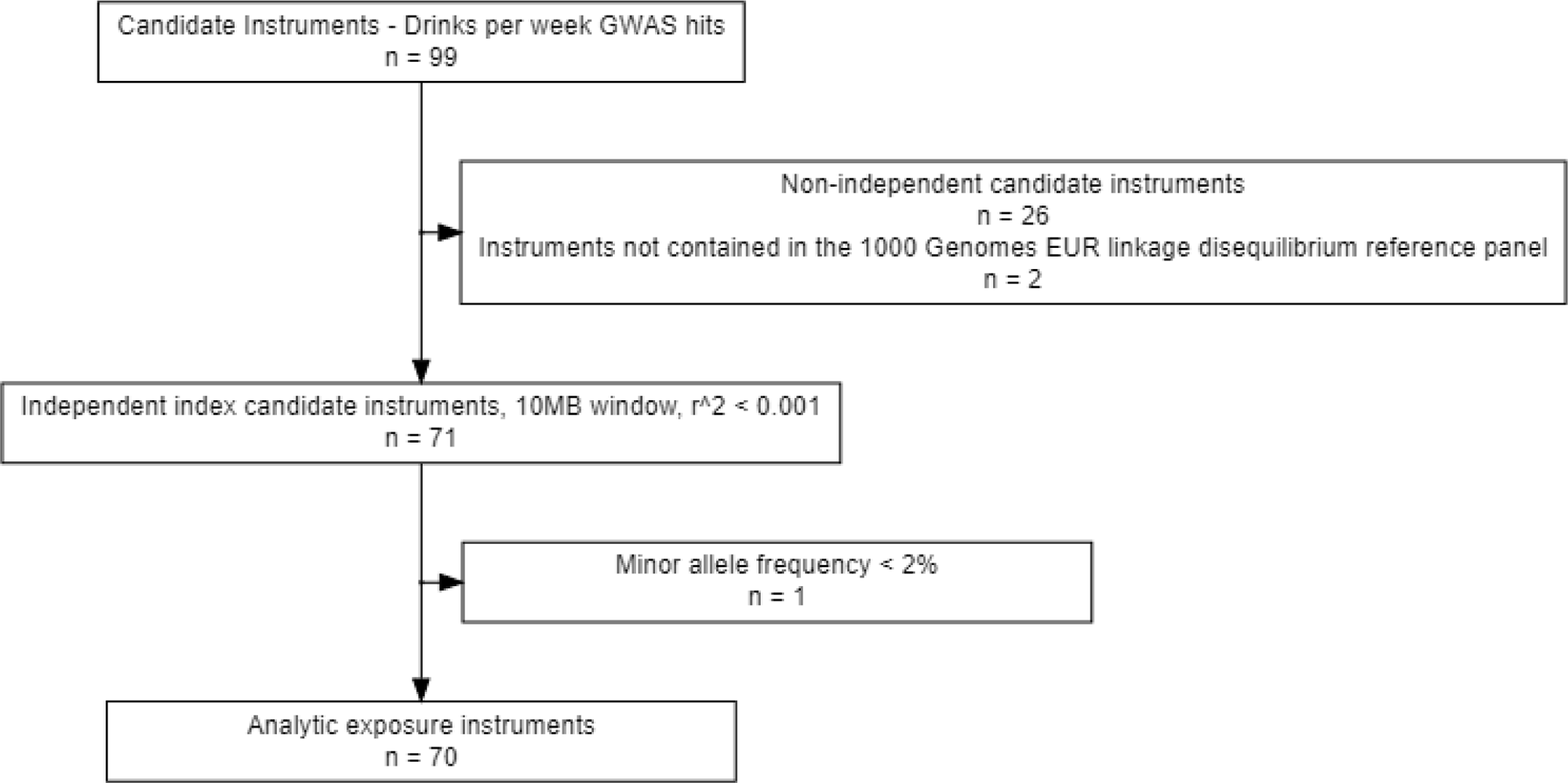
Inclusion-exclusion flowchart for one-sample Mendelian randomization single nucleotide polymorphism genetic instruments.

**Supplementary Figure 5.**
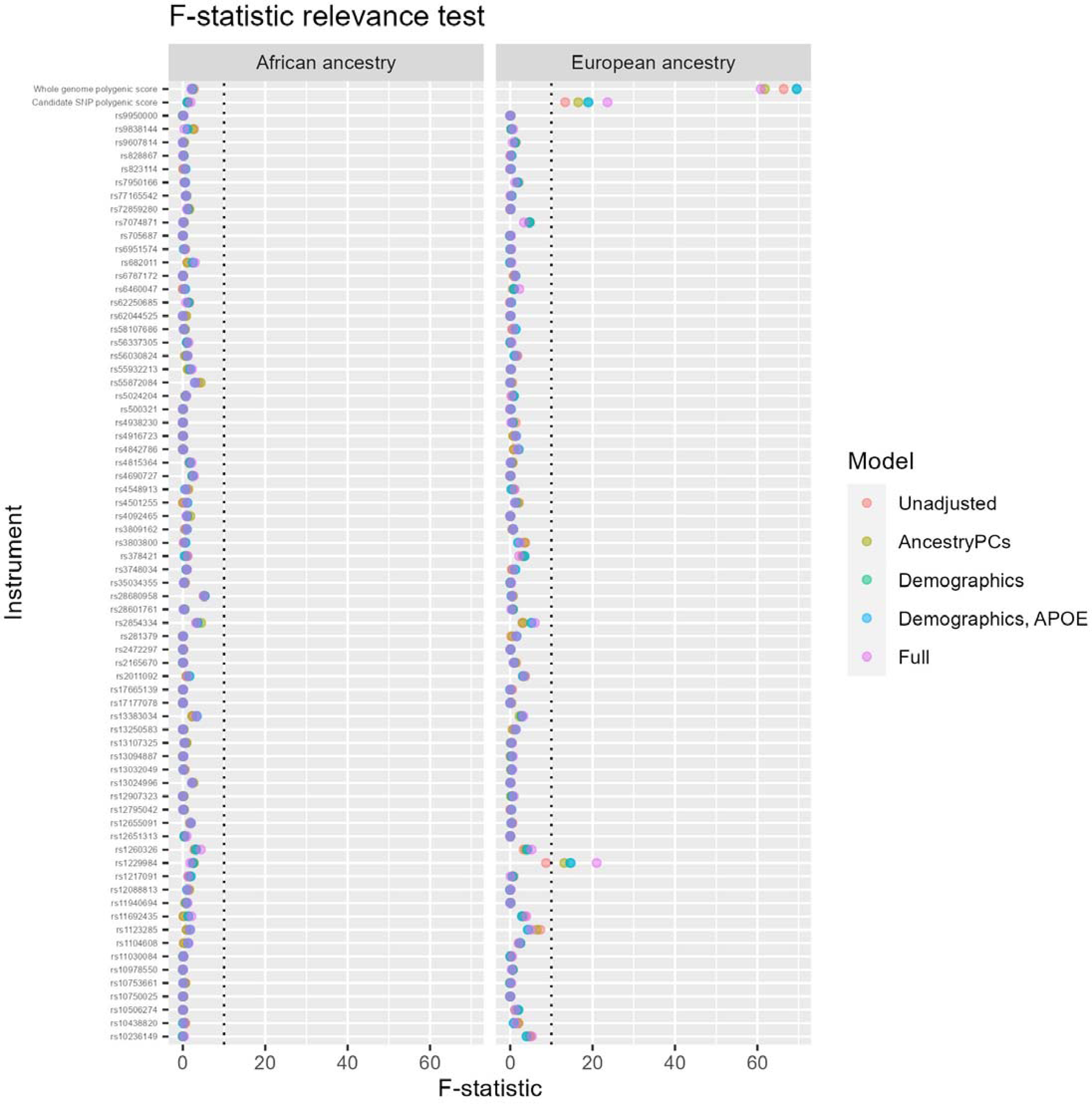
Relevance test of individual single nucleotide polymorphisms and polygenic risk scores.

**Supplementary Figure 6.**
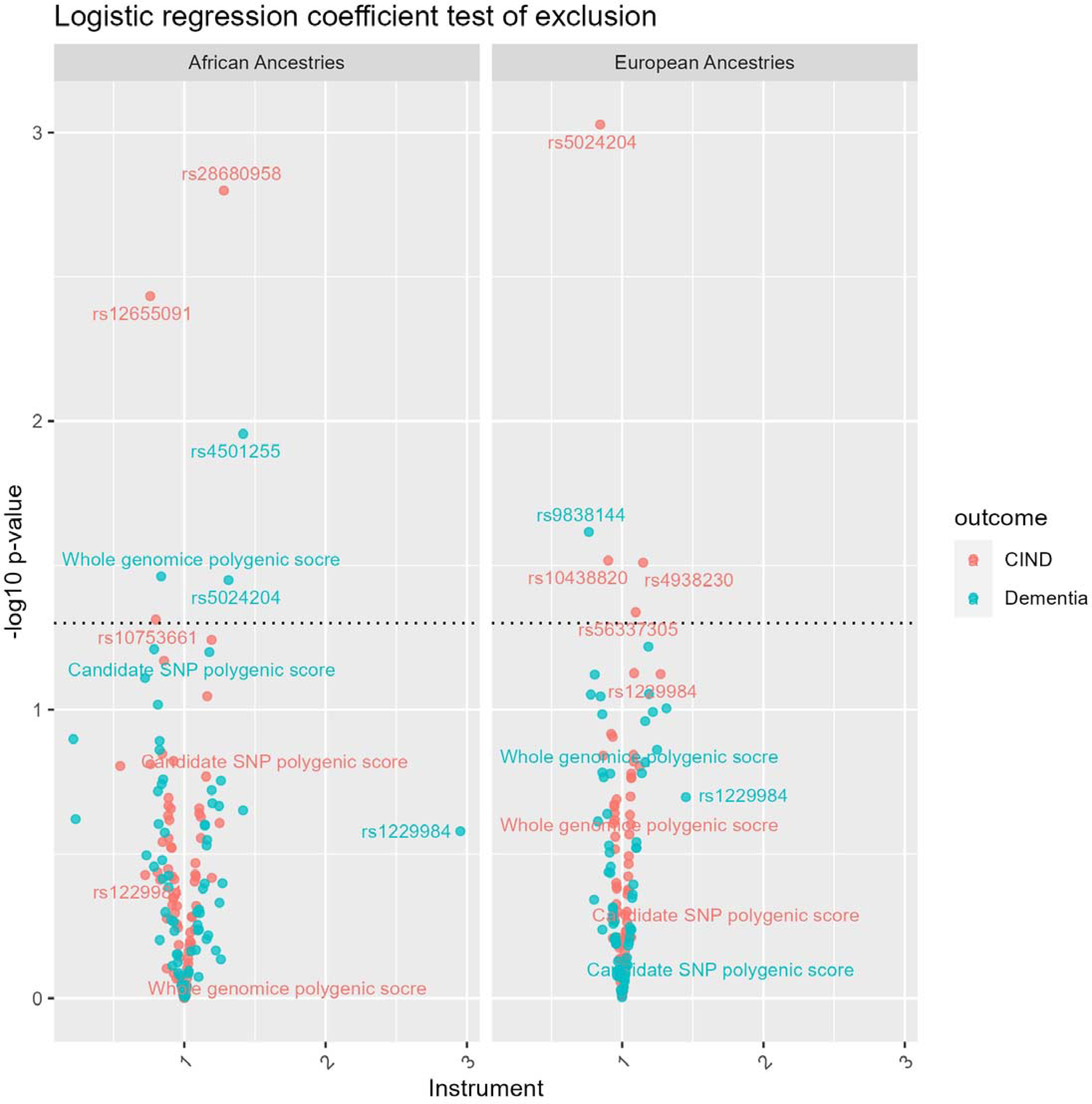
Exclusion test of individual SNPs and polygenic risk scores. The horizontal dotted line corresponds to a nominal p-value cutoff of 0.05. No results were significant after false discovery p-value adjustment.

**Supplementary Figure 7.**
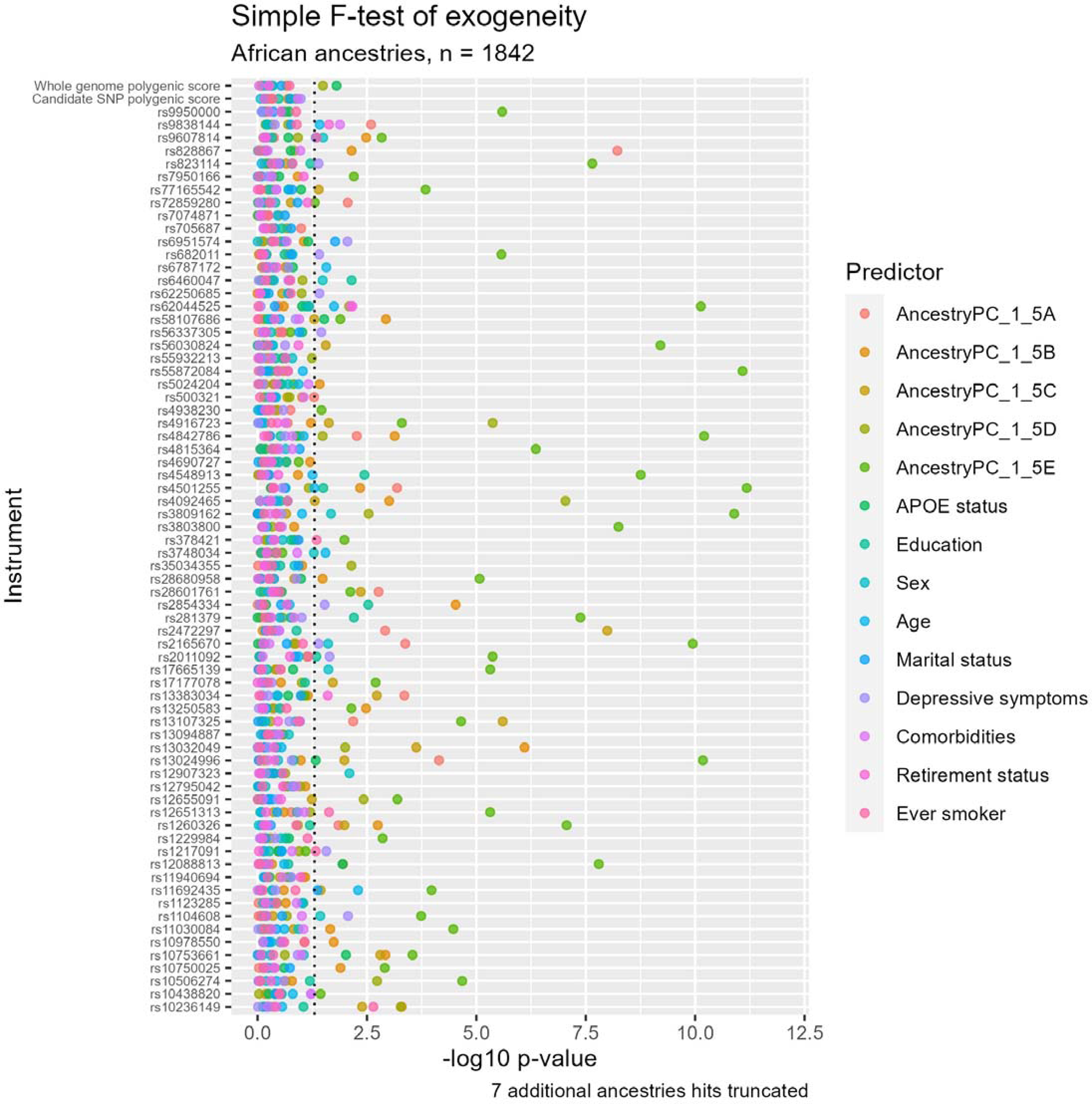
Exogeneity test of individual SNPs and polygenic risk scores in the African ancestries sample. The horizontal dotted line corresponds to a false discovery rate-adjusted p-value cutoff of 0.05.

**Supplementary Figure 8.**
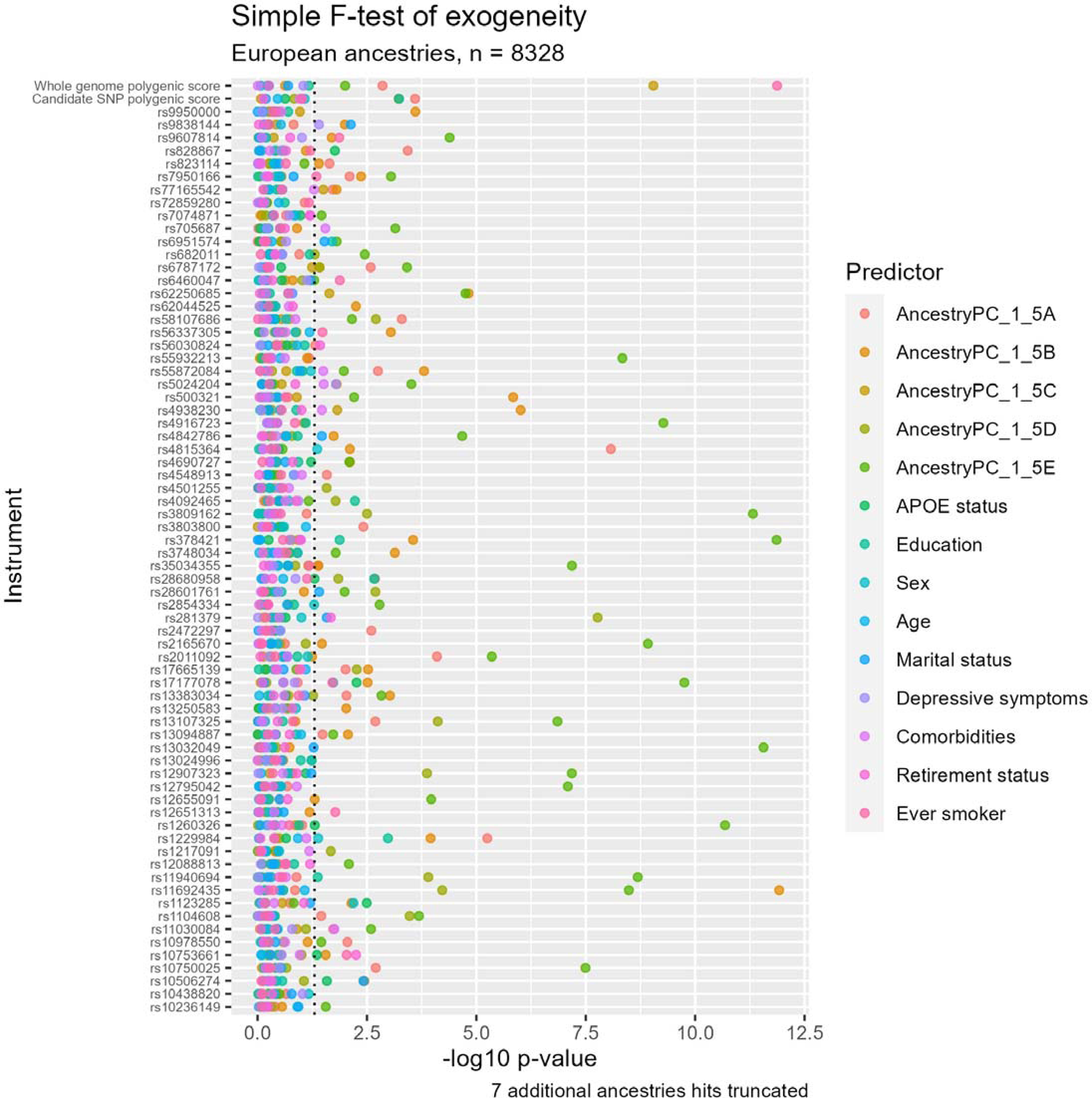
Exogeneity test of individual SNPs and polygenic risk scores in the European ancestries sample. The horizontal dotted line corresponds to a false discovery rate-adjusted p-value cutoff of 0.05.

**Supplementary Figure 9.**
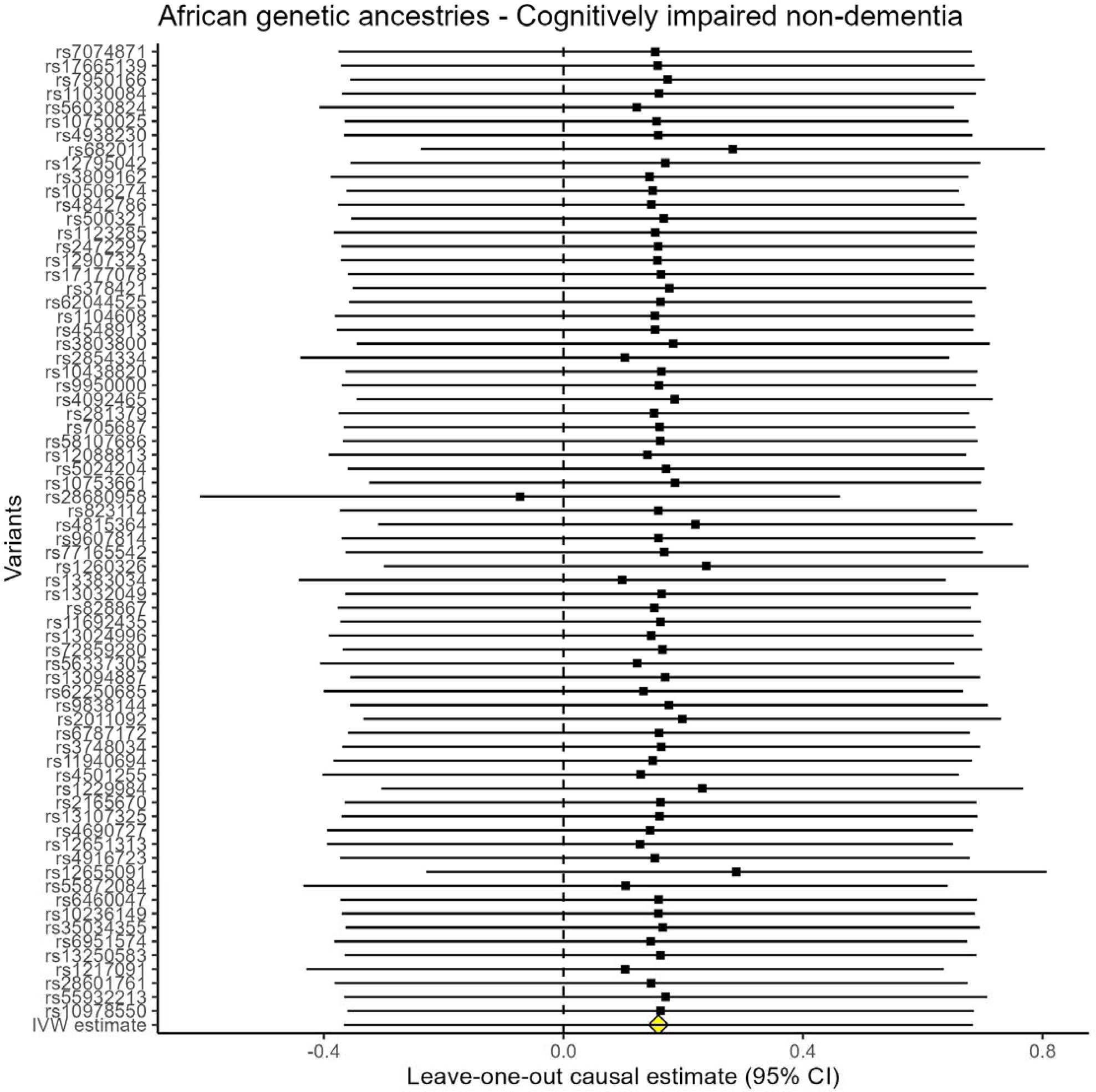
Leave-one-out sensitivity analysis for cognitively impaired non-dementia in the African genetic ancestries sample.

**Supplementary Figure 10.**
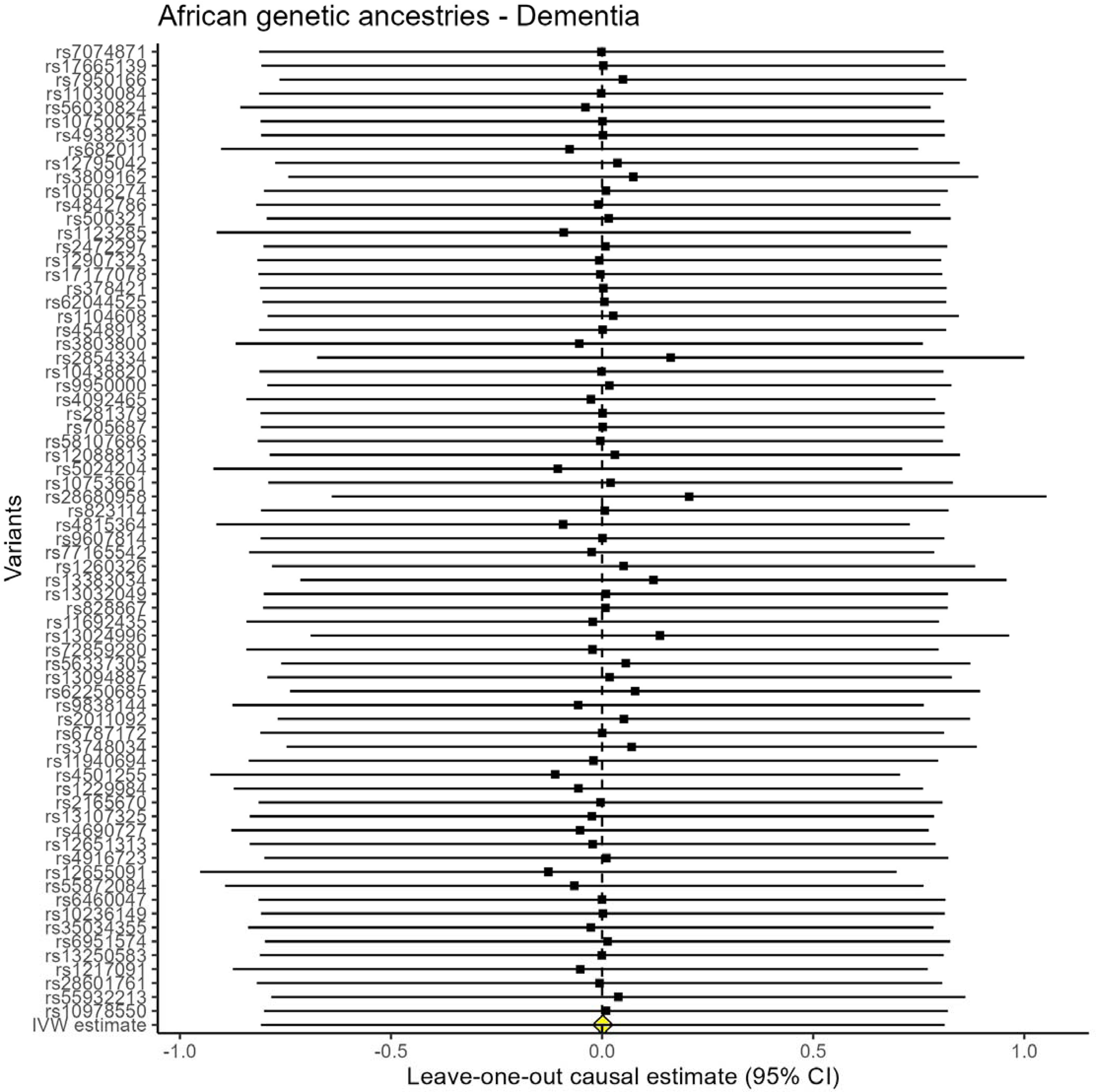
Leave-one-out sensitivity analysis for dementia in the African genetic ancestries sample.

**Supplementary Figure 11.**
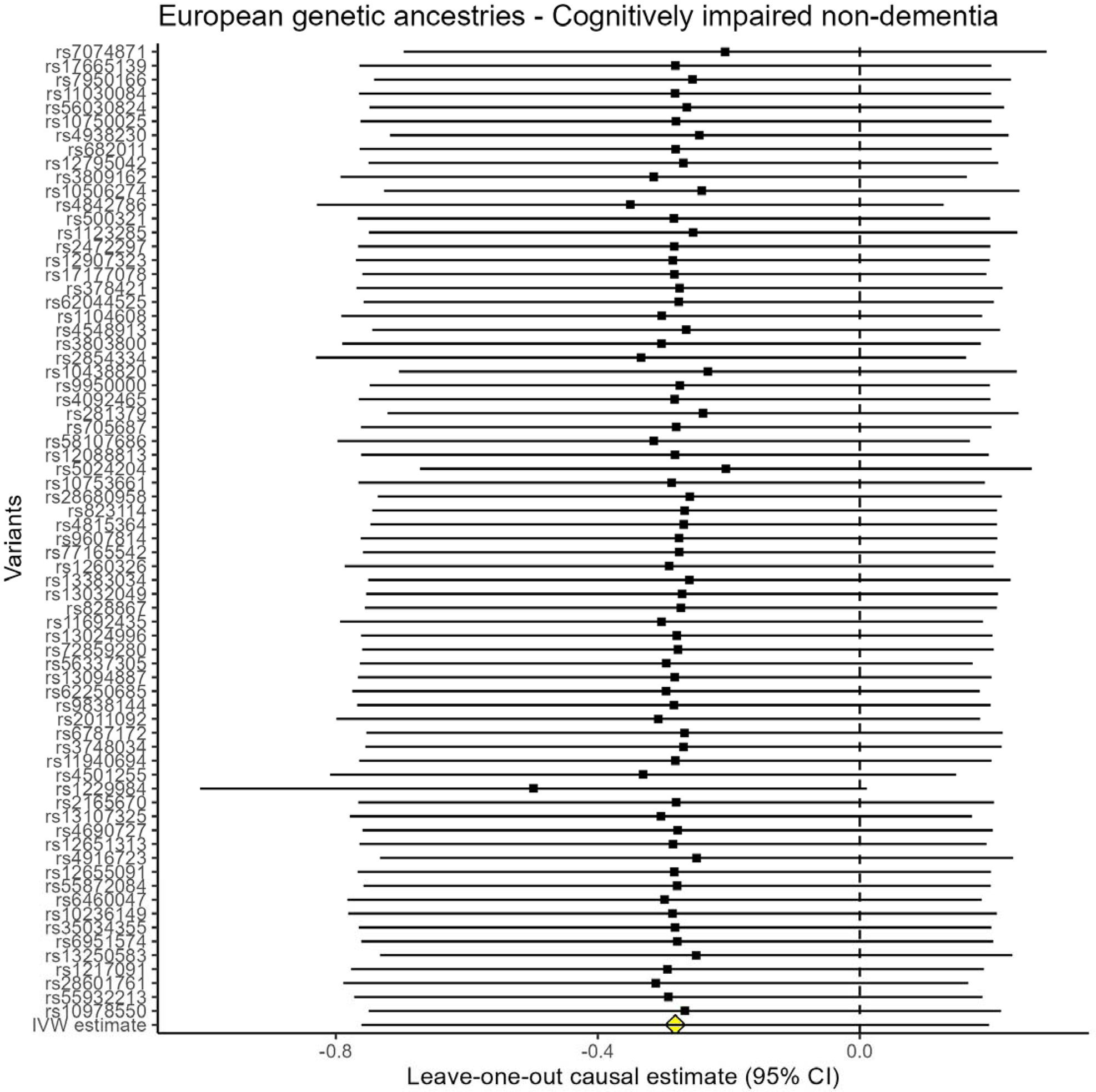
Leave-one-out sensitivity analysis for cognitively impaired non-dementia in the European genetic ancestries sample.

**Supplementary Figure 12.**
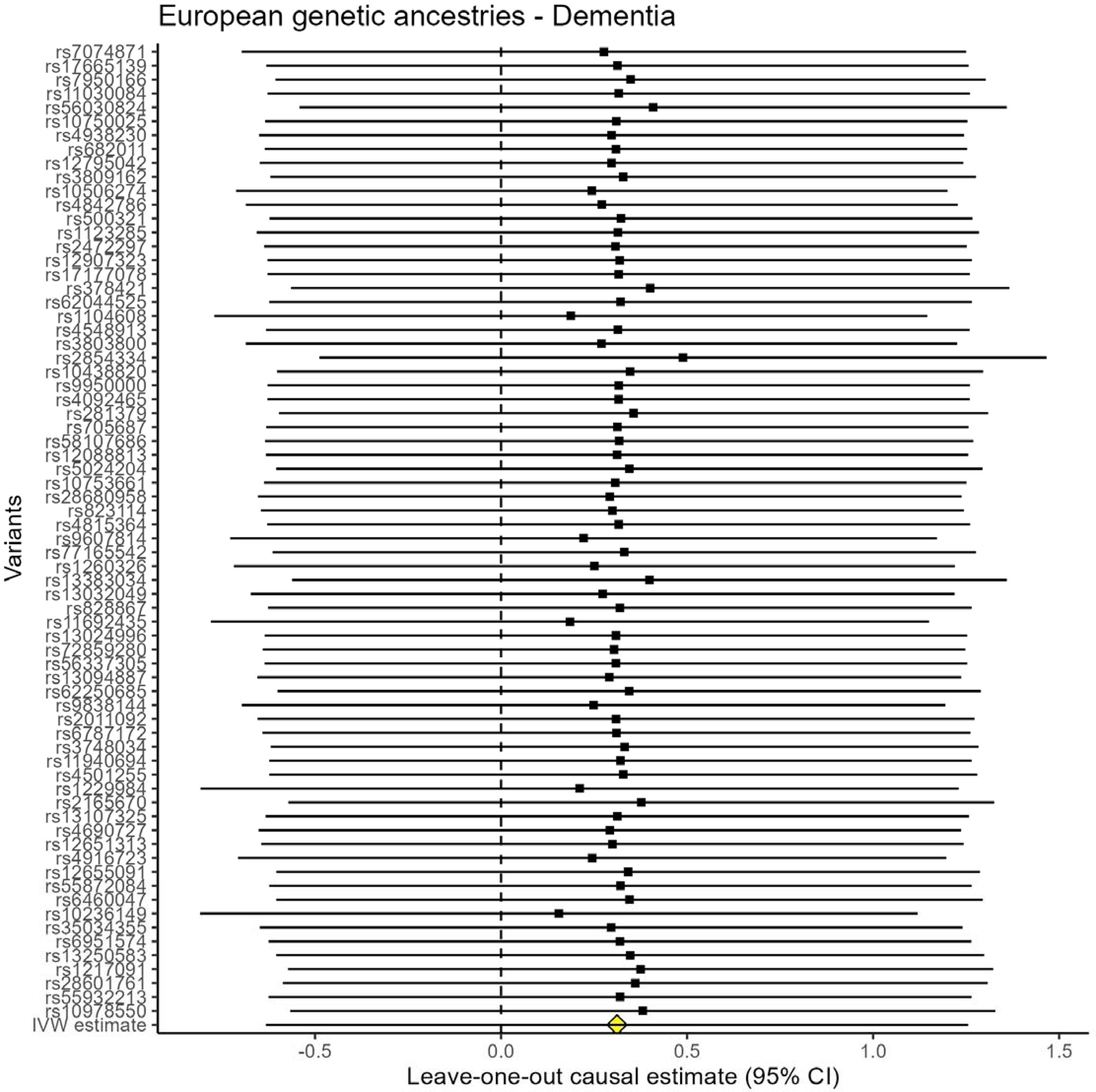
Leave-one-out sensitivity analysis for dementia in the European genetic ancestries sample.

**Supplementary Table 1.** Table of effect estimates of all conventional regression and Mendelian randomization tests for the African genetic ancestries sample at different covariate adjustment levels (n=1,842). *All Mendelian randomization models, including unadjusted models minimally adjusted for the first five principal components of genetic ancestry.

| African genetic ancestries |  |  |  |  |  |
| --- | --- | --- | --- | --- | --- |
| Method | Model* | Estimate | Lower 95% CI | Upper 95% CI | P-value |
| Dementia |  |  |  |  |  |
| Conventional regression | Unadjusted | 0.82 | 0.69 | 0.96 | 0.02 |
| Conventional regression | Age and sex | 0.54 | 0.45 | 0.64 | 0.00 |
| Conventional regression | Age, sex, educational attainment, APOE-ε4 allele status | 0.60 | 0.50 | 0.71 | 0.00 |
| Conventional regression | Fully adjusted | 0.98 | 0.82 | 1.17 | 0.86 |
| Whole-genome polygenic risk score (n=1,399,824 SNPs) for alcohol consumption | Unadjusted | 0.03 | 0.00 | 10.15 | 0.25 |
| Whole-genome polygenic risk score (n=1,399,824 SNPs) for alcohol consumption | Age and sex | 0.05 | 0.00 | 20.58 | 0.32 |
| Whole-genome polygenic risk score (n=1,399,824 SNPs) for alcohol consumption | Age, sex, educational attainment, APOE-ε4 allele status | 0.03 | 0.00 | 27.32 | 0.31 |
| Whole-genome polygenic risk score (n=1,399,824 SNPs) for alcohol consumption | Fully adjusted | 0.02 | 0.00 | 52.17 | 0.31 |
| Candidate instrumental variable (n=70 SNPs) polygenic risk score for alcohol consumption | Unadjusted | 131.25 | 0.01 | 1.84x10<sup>6</sup> | 0.32 |
| Candidate instrumental variable (n=70 SNPs) polygenic risk score for alcohol consumption | Age and sex | 1479.17 | 0.00 | 2.0x10<sup>9</sup> | 0.31 |
| Candidate instrumental variable (n=70 SNPs) polygenic risk score for alcohol consumption | Age, sex, educational attainment, APOE-ε4 allele status | 1434.85 | 0.00 | 2.8x10<sup>9</sup> | 0.32 |
| Candidate instrumental variable (n=70 SNPs) polygenic risk score for alcohol consumption | Fully adjusted | 95.09 | 0.03 | 3.6x10<sup>5</sup> | 0.28 |
| rs1229984 alone | Unadjusted | 24.37 | 0.14 | 4.2x10<sup>3</sup> | 0.22 |
| rs1229984 alone | Age and sex | 47.96 | 0.14 | 1.6x10<sup>4</sup> | 0.19 |
| rs1229984 alone | Age, sex, educational attainment, APOE-ε4 allele status | 249.88 | 0.10 | 6.1x10<sup>5</sup> | 0.17 |
| rs1229984 alone | Fully adjusted | 190.78 | 0.03 | 1.1x10<sup>6</sup> | 0.23 |
| Inverse variance weighted | Age and sex | 1.00 | 0.45 | 2.25 | 1.00 |
| Inverse variance weighted | Age, sex, educational attainment, APOE-ε4 allele status | 1.00 | 0.44 | 2.25 | 1.00 |
| Inverse variance weighted | Fully adjusted | 1.05 | 0.47 | 2.34 | 0.91 |
| Inverse variance weighted | Unadjusted | 0.88 | 0.40 | 1.90 | 0.74 |
| MR-Egger | Age and sex | 1.15 | 0.32 | 4.11 | 0.83 |
| MR-Egger | Age, sex, educational attainment, APOE-ε4 allele status | 1.28 | 0.36 | 4.53 | 0.70 |
| MR-Egger | Fully adjusted | 0.91 | 0.26 | 3.23 | 0.89 |
| MR-Egger | Unadjusted | 0.88 | 0.26 | 2.94 | 0.84 |
| MR-Egger intercept | Age and sex | 0.99 | 0.93 | 1.05 | 0.78 |
| MR-Egger intercept | Age, sex, educational attainment, APOE-ε4 allele status | 0.99 | 0.93 | 1.04 | 0.62 |
| MR-Egger intercept | Fully adjusted | 1.01 | 0.95 | 1.07 | 0.78 |
| MR-Egger intercept | Unadjusted | 1.00 | 0.94 | 1.06 | 0.99 |
| Simple median | Age and sex | 2.08 | 0.61 | 7.03 | 0.24 |
| Simple median | Age, sex, educational attainment, APOE-ε4 allele status | 2.55 | 0.76 | 8.62 | 0.13 |
| Simple median | Fully adjusted | 3.56 | 1.05 | 12.04 | 0.04 |
| Simple median | Unadjusted | 2.24 | 0.71 | 7.10 | 0.17 |
| Weighted median | Age and sex | 0.59 | 0.16 | 2.19 | 0.43 |
| Weighted median | Age, sex, educational attainment, APOE-ε4 allele status | 0.56 | 0.15 | 2.07 | 0.38 |
| Weighted median | Fully adjusted | 0.57 | 0.16 | 2.05 | 0.39 |
| Weighted median | Unadjusted | 0.45 | 0.13 | 1.56 | 0.21 |
| Cognitively impaired, non-dementia |  |  |  |  |  |
| Conventional regression | Unadjusted | 0.93 | 0.84 | 1.01 | 0.10 |
| Conventional regression | Age and sex | 0.82 | 0.77 | 0.86 | 0.00 |
| Method | Model* | Estimate | Lower 95% CI | Upper 95% CI | P-value |
| Conventional regression | Age, sex, educational attainment, APOE-ε4 allele status | 0.87 | 0.83 | 0.92 | 0.00 |
| Conventional regression | Fully adjusted | 0.96 | 0.86 | 1.06 | 0.41 |
| Whole-genome polygenic risk score (n=1,399,824 SNPs) for alcohol consumption | Unadjusted | 2.18 | 0.14 | 33.32 | 0.58 |
| Whole-genome polygenic risk score (n=1,399,824 SNPs) for alcohol consumption | Age and sex | 2.12 | 0.12 | 37.53 | 0.61 |
| Whole-genome polygenic risk score (n=1,399,824 SNPs) for alcohol consumption | Age, sex, educational attainment, APOE-ε4 allele status | 2.22 | 0.12 | 41.55 | 0.59 |
| Whole-genome polygenic risk score (n=1,399,824 SNPs) for alcohol consumption | Fully adjusted | 2.44 | 0.10 | 62.28 | 0.59 |
| Candidate instrumental variable (n=70 SNPs) polygenic risk score for alcohol consumption | Unadjusted | 0.06 | 0.00 | 15.68 | 0.33 |
| Candidate instrumental variable (n=70 SNPs) polygenic risk score for alcohol consumption | Age and sex | 0.05 | 0.00 | 30.33 | 0.36 |
| Candidate instrumental variable (n=70 SNPs) polygenic risk score for alcohol consumption | Age, sex, educational attainment, APOE-ε4 allele status | 0.03 | 0.00 | 54.94 | 0.35 |
| Candidate instrumental variable (n=70 SNPs) polygenic risk score for alcohol consumption | Fully adjusted | 0.04 | 0.00 | 8.18 | 0.24 |
| rs1229984 alone | Unadjusted | 0.30 | 0.03 | 3.29 | 0.32 |
| rs1229984 alone | Age and sex | 0.22 | 0.01 | 3.64 | 0.29 |
| rs1229984 alone | Age, sex, educational attainment, APOE-ε4 allele status | 0.21 | 0.01 | 5.97 | 0.36 |
| rs1229984 alone | Fully adjusted | 0.15 | 0.00 | 10.10 | 0.37 |
| Inverse variance weighted | Age and sex | 1.17 | 0.69 | 1.98 | 0.55 |
| Inverse variance weighted | Age, sex, educational attainment, APOE-ε4 allele status | 1.17 | 0.69 | 1.97 | 0.57 |
| Inverse variance weighted | Fully adjusted | 1.18 | 0.70 | 1.99 | 0.54 |
| Inverse variance weighted | Unadjusted | 1.08 | 0.66 | 1.79 | 0.76 |
| MR-Egger | Age and sex | 1.19 | 0.52 | 2.70 | 0.68 |
| MR-Egger | Age, sex, educational attainment, APOE-ε4 allele status | 1.13 | 0.50 | 2.54 | 0.77 |
| MR-Egger | Fully adjusted | 0.84 | 0.37 | 1.90 | 0.68 |
| MR-Egger | Unadjusted | 0.97 | 0.45 | 2.10 | 0.94 |
| MR-Egger intercept | Age and sex | 1.00 | 0.96 | 1.04 | 0.96 |
| MR-Egger intercept | Age, sex, educational attainment, APOE-ε4 allele status | 1.00 | 0.96 | 1.04 | 0.92 |
| MR-Egger intercept | Fully adjusted | 1.02 | 0.98 | 1.06 | 0.29 |
| MR-Egger intercept | Unadjusted | 1.01 | 0.97 | 1.05 | 0.71 |
| Simple median | Age and sex | 0.96 | 0.45 | 2.02 | 0.90 |
| Simple median | Age, sex, educational attainment, APOE-ε4 allele status | 0.87 | 0.41 | 1.85 | 0.72 |
| Simple median | Fully adjusted | 1.30 | 0.61 | 2.77 | 0.50 |
| Simple median | Unadjusted | 1.19 | 0.58 | 2.43 | 0.64 |
| Weighted median | Age and sex | 1.59 | 0.71 | 3.58 | 0.26 |
| Weighted median | Age, sex, educational attainment, APOE-ε4 allele status | 1.58 | 0.70 | 3.57 | 0.27 |
| Weighted median | Fully adjusted | 1.52 | 0.67 | 3.45 | 0.31 |
| Weighted median | Unadjusted | 1.56 | 0.71 | 3.41 | 0.27 |

**Supplementary Table 2.** Table of effect estimates of all conventional regression and Mendelian randomization tests for the European genetic ancestries sample at different covariate adjustment levels (n=8,328). *All Mendelian randomization models, including unadjusted models, minimally adjusted for the first five principal components of genetic ancestry.

| European genetic ancestries |  |  |  |  |  |
| --- | --- | --- | --- | --- | --- |
| Method | Model | Estimate | Lower 95% CI | Upper 95% CI | P-value |
| **Dementia** |  |  |  |  |  |
| Conventional regression | Unadjusted | 0.49 | 0.40 | 0.58 | 0.00 |
| Conventional regression | Age and sex | 0.54 | 0.45 | 0.64 | 0.00 |
| Conventional regression | Age, sex, educational attainment, APOE-ε4 allele status | 0.60 | 0.50 | 0.71 | 0.00 |
| Conventional regression | Fully adjusted | 0.62 | 0.51 | 0.73 | 0.00 |
| Whole-genome polygenic risk score (n=1,399,824 SNPs) for alcohol consumption | Unadjusted | 0.52 | 0.20 | 1.37 | 0.19 |
| Whole-genome polygenic risk score (n=1,399,824 SNPs) for alcohol consumption | Age and sex | 0.44 | 0.16 | 1.20 | 0.11 |
| Whole-genome polygenic risk score (n=1,399,824 SNPs) for alcohol consumption | Age, sex, educational attainment, APOE-ε4 allele status | 0.46 | 0.16 | 1.33 | 0.15 |
| Whole-genome polygenic risk score (n=1,399,824 SNPs) for alcohol consumption | Fully adjusted | 0.33 | 0.10 | 1.12 | 0.07 |
| Candidate instrumental variable (n=70 SNPs) polygenic risk score for alcohol consumption | Unadjusted | 0.46 | 0.06 | 3.36 | 0.44 |
| Candidate instrumental variable (n=70 SNPs) polygenic risk score for alcohol consumption | Age and sex | 0.44 | 0.06 | 3.49 | 0.44 |
| Candidate instrumental variable (n=70 SNPs) polygenic risk score for alcohol consumption | Age, sex, educational attainment, APOE-ε4 allele status | 0.38 | 0.05 | 3.04 | 0.36 |
| Candidate instrumental variable (n=70 SNPs) polygenic risk score for alcohol consumption | Fully adjusted | 0.29 | 0.04 | 2.37 | 0.25 |
| rs1229984 alone | Unadjusted | 2.55 | 0.14 | 48.26 | 0.53 |
| rs1229984 alone | Age and sex | 2.50 | 0.14 | 45.59 | 0.54 |
| rs1229984 alone | Age, sex, educational attainment, APOE-ε4 allele status | 2.33 | 0.13 | 42.93 | 0.57 |
| rs1229984 alone | Fully adjusted | 1.69 | 0.12 | 24.24 | 0.70 |
| Inverse variance weighted | Age and sex | 1.37 | 0.53 | 3.51 | 0.52 |
| Inverse variance weighted | Age, sex, educational attainment, APOE-ε4 allele status | 1.39 | 0.54 | 3.57 | 0.50 |
| Inverse variance weighted | Fully adjusted | 1.67 | 0.66 | 4.24 | 0.28 |
| Inverse variance weighted | Unadjusted | 1.15 | 0.45 | 2.92 | 0.77 |
| MR-Egger | Age and sex | 2.29 | 0.56 | 9.37 | 0.25 |
| MR-Egger | Age, sex, educational attainment, APOE-ε4 allele status | 2.26 | 0.56 | 9.09 | 0.25 |
| MR-Egger | Fully adjusted | 1.48 | 0.39 | 5.63 | 0.57 |
| MR-Egger | Unadjusted | 2.07 | 0.52 | 8.21 | 0.30 |
| MR-Egger intercept | Age and sex | 0.98 | 0.95 | 1.02 | 0.33 |
| MR-Egger intercept | Age, sex, educational attainment, APOE-ε4 allele status | 0.98 | 0.95 | 1.02 | 0.35 |
| MR-Egger intercept | Fully adjusted | 1.00 | 0.97 | 1.04 | 0.80 |
| MR-Egger intercept | Unadjusted | 0.98 | 0.95 | 1.02 | 0.26 |
| Simple median | Age and sex | 0.45 | 0.11 | 1.88 | 0.27 |
| Simple median | Age, sex, educational attainment, APOE-ε4 allele status | 0.85 | 0.20 | 3.67 | 0.83 |
| Simple median | Fully adjusted | 1.37 | 0.32 | 5.91 | 0.68 |
| Simple median | Unadjusted | 0.45 | 0.11 | 1.87 | 0.27 |
| Weighted median | Age and sex | 1.89 | 0.44 | 8.20 | 0.40 |
| Weighted median | Age, sex, educational attainment, APOE-ε4 allele status | 1.82 | 0.40 | 8.21 | 0.44 |
| Weighted median | Fully adjusted | 1.87 | 0.41 | 8.55 | 0.42 |
| Weighted median | Unadjusted | 1.72 | 0.41 | 7.27 | 0.46 |
| **Cognitively impaired, non-dementia** |  |  |  |  |  |
| Conventional regression | Unadjusted | 0.78 | 0.74 | 0.82 | 0.00 |
| Conventional regression | Age and sex | 0.82 | 0.77 | 0.86 | 0.00 |
| Method | Model | Estimate | Lower 95% CI | Upper 95% CI | P-value |
| Conventional regression | Age, sex, educational attainment, APOE-ε4 allele status | 0.87 | 0.83 | 0.92 | 0.00 |
| Conventional regression | Fully adjusted | 0.89 | 0.84 | 0.94 | 0.00 |
| Whole-genome polygenic risk score (n=1,399,824 SNPs) for alcohol consumption | Unadjusted | 0.71 | 0.43 | 1.18 | 0.18 |
| Whole-genome polygenic risk score (n=1,399,824 SNPs) for alcohol consumption | Age and sex | 0.69 | 0.41 | 1.15 | 0.16 |
| Whole-genome polygenic risk score (n=1,399,824 SNPs) for alcohol consumption | Age, sex, educational attainment, APOE-ε4 allele status | 0.66 | 0.39 | 1.11 | 0.12 |
| Whole-genome polygenic risk score (n=1,399,824 SNPs) for alcohol consumption | Fully adjusted | 0.57 | 0.31 | 1.05 | 0.07 |
| Candidate instrumental variable (n=70 SNPs) polygenic risk score for alcohol consumption | Unadjusted | 1.22 | 0.43 | 3.44 | 0.71 |
| Candidate instrumental variable (n=70 SNPs) polygenic risk score for alcohol consumption | Age and sex | 1.44 | 0.51 | 4.07 | 0.50 |
| Candidate instrumental variable (n=70 SNPs) polygenic risk score for alcohol consumption | Age, sex, educational attainment, APOE-ε4 allele status | 1.22 | 0.46 | 3.23 | 0.69 |
| Candidate instrumental variable (n=70 SNPs) polygenic risk score for alcohol consumption | Fully adjusted | 1.14 | 0.43 | 3.07 | 0.79 |
| rs1229984 alone | Unadjusted | 2.47 | 0.67 | 9.12 | 0.17 |
| rs1229984 alone | Age and sex | 3.31 | 0.87 | 12.54 | 0.08 |
| rs1229984 alone | Age, sex, educational attainment, APOE-ε4 allele status | 3.36 | 0.95 | 11.86 | 0.06 |
| rs1229984 alone | Fully adjusted | 2.79 | 0.90 | 8.68 | 0.08 |
| Inverse variance weighted | Age and sex | 0.75 | 0.47 | 1.22 | 0.25 |
| Inverse variance weighted | Age, sex, educational attainment, APOE-ε4 allele status | 0.80 | 0.50 | 1.30 | 0.37 |
| Inverse variance weighted | Fully adjusted | 0.86 | 0.53 | 1.37 | 0.52 |
| Inverse variance weighted | Unadjusted | 0.73 | 0.46 | 1.18 | 0.20 |
| MR-Egger | Age and sex | 1.10 | 0.54 | 2.22 | 0.80 |
| MR-Egger | Age, sex, educational attainment, APOE-ε4 allele status | 1.14 | 0.56 | 2.29 | 0.72 |
| MR-Egger | Fully adjusted | 1.04 | 0.53 | 2.05 | 0.91 |
| MR-Egger | Unadjusted | 1.11 | 0.56 | 2.20 | 0.77 |
| MR-Egger intercept | Age and sex | 0.99 | 0.97 | 1.01 | 0.16 |
| MR-Egger intercept | Age, sex, educational attainment, APOE-ε4 allele status | 0.99 | 0.97 | 1.01 | 0.19 |
| MR-Egger intercept | Fully adjusted | 0.99 | 0.98 | 1.01 | 0.43 |
| MR-Egger intercept | Unadjusted | 0.99 | 0.97 | 1.00 | 0.11 |
| Simple median | Age and sex | 0.58 | 0.28 | 1.20 | 0.14 |
| Simple median | Age, sex, educational attainment, APOE-ε4 allele status | 0.61 | 0.29 | 1.30 | 0.20 |
| Simple median | Fully adjusted | 0.73 | 0.34 | 1.56 | 0.42 |
| Simple median | Unadjusted | 0.59 | 0.29 | 1.23 | 0.16 |
| Weighted median | Age and sex | 0.90 | 0.44 | 1.84 | 0.77 |
| Weighted median | Age, sex, educational attainment, APOE-ε4 allele status | 0.93 | 0.45 | 1.91 | 0.84 |
| Weighted median | Fully adjusted | 1.25 | 0.63 | 2.51 | 0.52 |
| Weighted median | Unadjusted | 0.88 | 0.43 | 1.79 | 0.73 |

